# Mathematical modeling of COVID-19 in British Columbia: an age-structured model with time-dependent contact rates

**DOI:** 10.1101/2021.10.19.21265177

**Authors:** Sarafa A. Iyaniwura, Rebeca C. Falcão, Notice Ringa, Prince A. Adu, Michelle Spencer, Marsha Taylor, Caroline Colijn, Daniel Coombs, Naveed Z. Janjua, Michael A Irvine, Michael Otterstatter

**Affiliations:** British Columbia Centre for Disease Control, Vancouver, British Columbia, Canada; Department of Mathematics and Institute of Applied Mathematics, University of British Columbia, Vancouver, British Columbia, Canada; Department of Mathematics and Statistics, Simon Fraser University, Burnaby, BC, Canada; School of Population and Public Health, University of British Columbia, Vancouver, British Columbia, Canada; Faculty of Health Sciences, Simon Fraser University, Burnaby, British Columbia, Canada

**Keywords:** SARS-CoV-2, COVID-19, age-structured model, mixing patterns, epidemics, Bayesian inference

## Abstract

Following the emergence of COVID-19 at the end of 2019, several mathematical models have been developed to study the transmission dynamics of this disease. Many of these models assume homogeneous mixing in the underlying population. However, contact rates and mixing patterns can vary dramatically among individuals depending on their age and activity level. Variation in contact rates among age groups and over time can significantly impact how well a model captures observed trends. To properly model the age-dependent dynamics of COVID-19 and understand the impacts of interventions, it is essential to consider heterogeneity arising from contact rates and mixing patterns. We developed an age-structured model that incorporates time-varying contact rates and population mixing computed from the ongoing BC Mix COVID-19 survey to study transmission dynamics of COVID-19 in British Columbia (BC), Canada. Using a Bayesian inference framework, we fit four versions of our model to weekly reported cases of COVID-19 in BC, with each version allowing different assumptions of contact rates. We show that in addition to incorporating age-specific contact rates and mixing patterns, time-dependent (weekly) contact rates are needed to adequately capture the observed transmission dynamics of COVID-19. Our approach provides a framework for explicitly including empirical contact rates in a transmission model, which removes the need to otherwise model the impact of many non-pharmaceutical interventions. Further, this approach allows projection of future cases based on clear assumptions of age-specific contact rates, as opposed to less tractable assumptions regarding transmission rates.

## 1 Introduction

COVID-19 disease is an infectious respiratory disease caused by the severe acute respiratory syndrome coronavirus 2 (SARS-CoV-2) [2]. It was first detected in the city of Wuhan, Hubei province, China, in December 2019 [65] and has since spread around the world with over 235 million confirmed cases and over 4.8 million confirmed deaths globally as of October 3, 2021 [64]. The World Health Organization (WHO) declared COVID-19 a public health emergency of international concern (PHEIC) on January 20, 2020 [66] and a pandemic on March 11, 2020 [67]. Several non-pharmaceutical interventions (NPIs), such as physical distancing, isolation, hand washing, stay-at-home orders, closure of schools and businesses, and travel restrictions, have been deployed at various times to limit the spread of COVID-19 [22, 23, 42, 49]. Nevertheless, significant numbers of new COVID-19 cases continue to occur worldwide.

The first laboratory-confirmed case of COVID-19 infection in the province of British Columbia (BC), Canada, was reported to public health on January 26, 2020 [51]. As of October 3, 2021, there have been over 189,000 reported cases of COVID-19 in BC and nearly 2,000 confirmed deaths [26]. The distribution of reported cases varies between age groups, with generally a small proportion among children and a declining proportion with age among adults [25]. However, the proportion of cases hospitalized generally increases with age. In addition to other factors, including public health measures, these differences may be due to underlying variation in contact rates and activity by age, as well variation in susceptibility [17, 7, 69] and severity [38, 59, 33, 19] by age. To provide a better understanding of the transmission dynamics of COVID-19, it is therefore essential to assess the impact of contact rates and mixing patterns of individuals among different age groups [12, 45, 35, 50, 69, 54].

Several mathematical models have been deployed to study the transmission dynamics of COVID-19. Many of these models assume homogeneous mixing in the population, using the same contact rate and mixing pattern for everyone [3, 30, 58, 8, 48, 41, 9, 34]. These types of models can help to answer broad population-level questions, but are not suitable for understanding age-related dynamics. Age-structured models have also been used to study COVID-19, including the impact of social distancing on its spread [39, 54], the effectiveness of different vaccination strategies [46, 47, 13, 40, 53, 29, 52, 36], and the effects of herd immunity and re-opening [12, 11, 10, 62, 14, 5]. Although these models incorporate heterogeneity in the contact rates and mixing patterns by age group, they assume fixed contact rate for each age group throughout the epidemic. This can be problematic since NPIs, such as physical distancing, self-isolation, stay-at-home orders, and travel restrictions alter contact and activity levels over time, and these impacts may differ by age group. In order to overcome these limitations, it is necessary to incorporate ongoing population-based measures of contact rates by age group into a transmission modeling approach.

In this work, we develop a susceptible-exposed-infected-recovered (SEIR) age-structured model to study the transmission dynamics of COVID-19 in British Columbia, Canada. This model incorporates the average weekly contact rates and population mixing patterns computed from the ongoing BC Mix COVID-19 Survey (BC-Mix) [1]. Unlike other age-structured models used to study COVID-19 dynamics, where contact rate for each age group is fixed throughout the epidemic or adjusted by re-scaling an initial rate, our model uses the near real-time contact rates among age groups to inform transmission dynamics. Using a Bayesian inference framework, we calibrate our model to the weekly reported cases of COVID-19 in BC. Scaling parameters are estimated by age group to account for the differential impact of contacts on transmission. We considered four different scenarios that allow or exclude variation in contact rates and scaling parameters in our model fitting and show that optimal fits require both the age-specific scaling and time-dependent contact rates.

## 2 Methods

### 2.1 Mathematical Model

We develop an age-structured SEIR model to study the transmission dynamics of COVID-19 in British Columbia (BC), Canada. Our model follows a design similar to an age- and activity-structured model used previously to study the dynamics of the 2009 H1N1 influenza pandemic in the Greater Vancouver Regional District of British Columbia [16]. It has six compartments tracking the disease trajectory: where *S* represents the susceptible population, *E*_1_ and *E*_2_ are for the exposed population, *I*_1_ and *I*_2_ represent the infected population, and *R* represents those who have recovered from the disease. We split the exposed and infected compartments into two so that the time an individual spends in these states follows a gamma distribution, which is more realistic than an exponential distribution. As well, individuals in *E*_1_ are considered exposed, but not yet able to transmit infection (exposed), while those in *E*_2_ can transmit infections without showing any symptoms of the disease (pre-symptomatic). An infected individual spends the first half of their infectious period in *I*_1_ and the other half in *I*_2_. Here, we used an infectious period of 5 days [57, 27, 3]. See Table 1 for all the model parameters. Figure 1 shows the schematic diagram of the model with solid black arrows indicating the direction of flow of individuals between the compartments at the rates indicated beside the arrows, while the dashed red arrows show disease transmission. The index *j* represents the age groups. The compartments *E*_2_, *I*_1_, and *I*_2_ (in red) are the infectious compartments of the model, representing individuals who may transmit the disease to their contacts. We assume that the total population size of BC remains constant over the epidemic period and that recovered individuals develop permanent immunity to the virus.

**Table 1:** Model parameters, descriptions, and values. *The basic reproduction number R*_0_ *in the SEIR model* (2.1) *is taken to be the effective reproduction number R*_0*t*_ *in BC at the beginning of September 2020 since the case data considered are from September 2020 to January 2021. The normalization constant C*_*N*_ *for R*_0*t*_ *was computed for the two population mixing matrices shown in Figure 3. For the BC Mix matrix (left panel of Figure 3) C*_*N*_ = 35.1027 *and for the 2009 matrix (right panel of Figure 3) C*_*N*_ = 30.2650. *The total population of BC is 5,147,712 [6]*.

| Parameter | Description | Value | References |
| --- | --- | --- | --- |
| $N_{\text{age}}$ | Number of age groups | 10 | |
| $\beta$ | Disease transmission rate | | Derived [1] |
| $C$ | $N_{\text{age}} \times N_{\text{age}}$ population mixing matrix | | Derived [1] |
| $K$ | Contact rate vector | | Derived [1] |
| $\Phi$ | Vector of the scaling parameters for the contact rates | | Estimated |
| $H_1$ | Rate of transitioning from $E_1$ to $E_2$ | 1/5 (days<sup>-1</sup>) | [41, 70, 3] |
| $H_2$ | Rate of transitioning from $E_2$ to $I_1$ | 1 (days<sup>-1</sup>) | [43, 57, 27, 3] |
| $\gamma$ | Infection recovery rate | 1/5 (days<sup>-1</sup>) | [57, 27, 3] |
| $R_{0t}$ | Effective reproduction number in BC at the beginning of September 2020 | 1.24 | [24] |
| $C_N$ | Normalization constant for $R_{0t}$ | 35.1027 & 30.2650 | Computed |
| $P_{\text{age}}$ | Population of British Columbia by age-group | 86,903 (< 2 years) | [6] |
|  |  | 187,311 (2 – 5 years) |  |
|  |  | 599,526 (6 – 17 years) |  |
|  |  | 458,056 (18 – 24 years) |  |
|  |  | 734,923 (25 – 34 years) |  |
|  |  | 687,491 (35 – 44 years) |  |
|  |  | 669,568 (45 – 54 years) |  |
|  |  | 736,998 (55 – 64 years) |  |
|  |  | 576,218 (65 – 74 years) |  |
|  |  | 410,718 (75+ years) |  |

**Figure 1:**
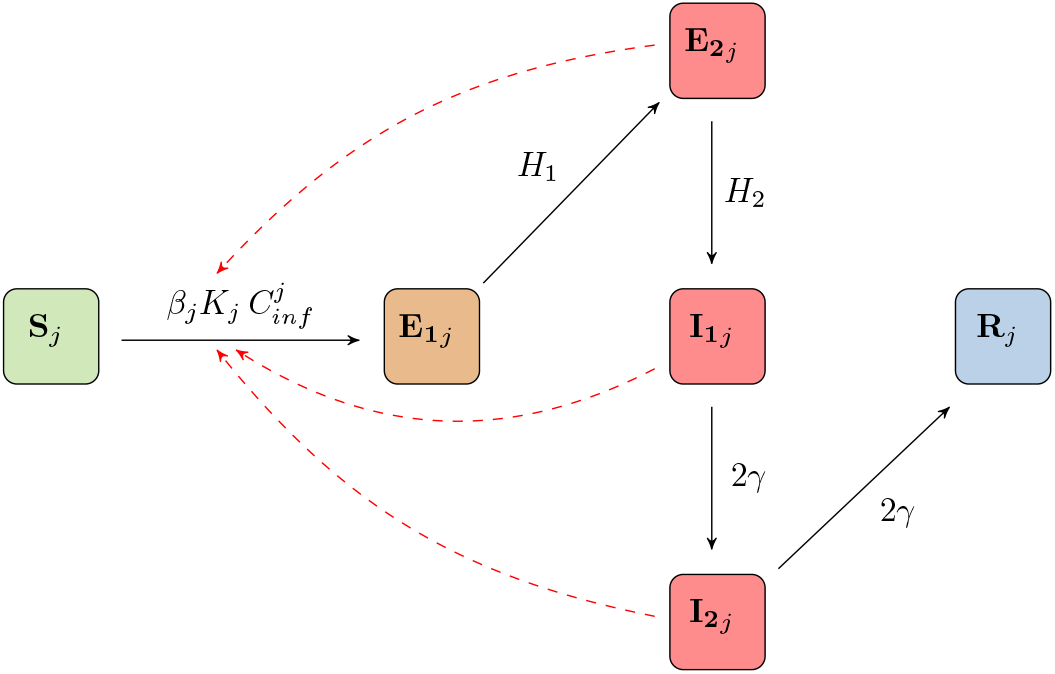
Schematic of the age-structured model. *Compartments are defined as follows: Susceptible individuals (S); exposed (E*_1_*); pre-symptomatic (E*_2_*); Infected individuals in the first half of their infectious period (I*_1_*); Infected individuals in the second half of their infectious period (I*_2_*); and recovered (R). It is assumed that recovered individuals become immune to the disease. Each compartment is a vector whose elements represent the corresponding population for each age-group in the model. The index in the schematic represents the j*^*th*^ *age group. The solid black arrows show the flow of individuals between the model compartments at rates indicated beside the arrows, while the dashed red arrows indicated disease transmission (see* (2.1) *for more details)*.

Let *N*_age_ be the number of age groups in the model so that each of the compartments *S, E*_1_, *E*_2_, *I*_1_, *I*_2_, and *R* is a vector with *N*_age_ elements. We stratify the BC population into *N*_*age*_ = 10 age groups given by *<* 2 years, 2-5 years, 6-17 years, 18-24 years, 25-34 years, 35-44 years, 45-54 years, 55-64 years, 65-74 years, and 75+ years. These age groups largely follow those used in the influenza study of [16]; however, other groupings could be used within this framework. Each compartment of the model is divided into ten sub-compartments based on age groups. This gives a total of 60 sub-compartments and ordinary differential equations for the model. Our model further assumes that each individual remains in the same age group throughout the epidemic. The differential equations for the model in vector notation are given by:

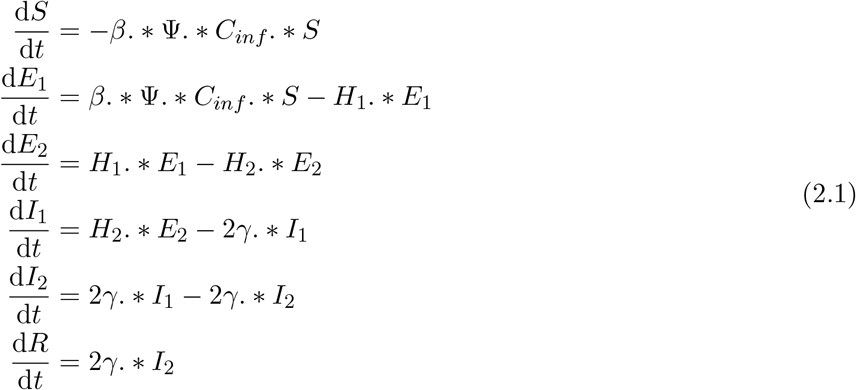

where *H*_1_ is the rate of transitioning from *E*_1_ to *E*_2_, *H*_2_ is the rate of transitioning from *E*_2_ to *I*_1_, and *γ* is the recovery rate. We have assumed that the transition rates *H*_1_, *H*_2_, and *γ* are the same for all age groups. The model can easily be extended to have age-specific transition rates. In this case, *H*_1_, *H*_2_, and *γ* would be vectors with *N*_age_ elements. Note that. *\** represent element-wise multiplication. The vector Ψ = Φ. *\* K*, where *K* is a vector for the contact rates of the age groups and Φ ≡ (*ϕ*_1_, …, *ϕ*_Nage_) is a vector whose entries are the scaling parameters for the contact rates. These scaling parameters are used to account for the differential impact of contacts on transmission and are fixed throughout the epidemic. We consider a scenario where the contact rate *K* is fixed throughout the epidemic period and another where it changes from week to week during the epidemic. In addition, a fixed scaling parameter can be used for the entire population; in this case, Φ becomes a scalar. More details about the scaling parameters and the different scenarios can be found in Section 2.3.

In the ODE model (2.1), *β* is the transmission rate given by

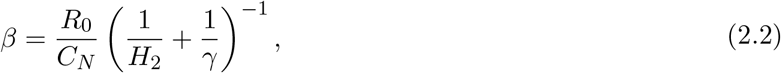

where *R*_0_ is the basic reproduction number and *C*_*N*_ is the normalization constant for *R*_0_ that incorporates the effect of the contact rates and the population mixing patterns into the basic reproduction number. Details of how *C*_*N*_ is calculated are given below. The fractions 1*/H*_2_ and 1*/γ* are the pre-symptomatic and recovery periods, respectively. The heterogeneity in disease transmission depends largely on the vector *C*_*inf*_, whose *j*^th^ entry is defined as

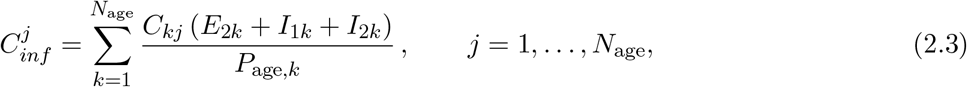

where *C*_*kj*_ is the *k*^th^ element in the *j*^th^ column of the *N*_age_ *× N*_age_ mixing matrix *C*, which gives the proportion of the total contacts of the *j*^th^ age group that is made with the *k*^th^ age group (see Figure 3). Here, *P*_age_ is a vector for the BC population size stratified by age group and *P*_age,*k*_ is the population of the *k*^th^ age group. The expression (*E*_2*k*_ + *I*_1*k*_ + *I*_2*k*_)*/P*_age,*k*_ gives the proportion of infectious individuals in the *k*^th^ age group. Unlike the model of [16], we do not further stratify our population by activity levels here. The activity level of each age group is implicitly captured by their average weekly contact rates. We computed the average weekly contact rates *K* and population mixing matrix *C* from the BC Mix COVID-19 survey [1].

**Figure 2:**
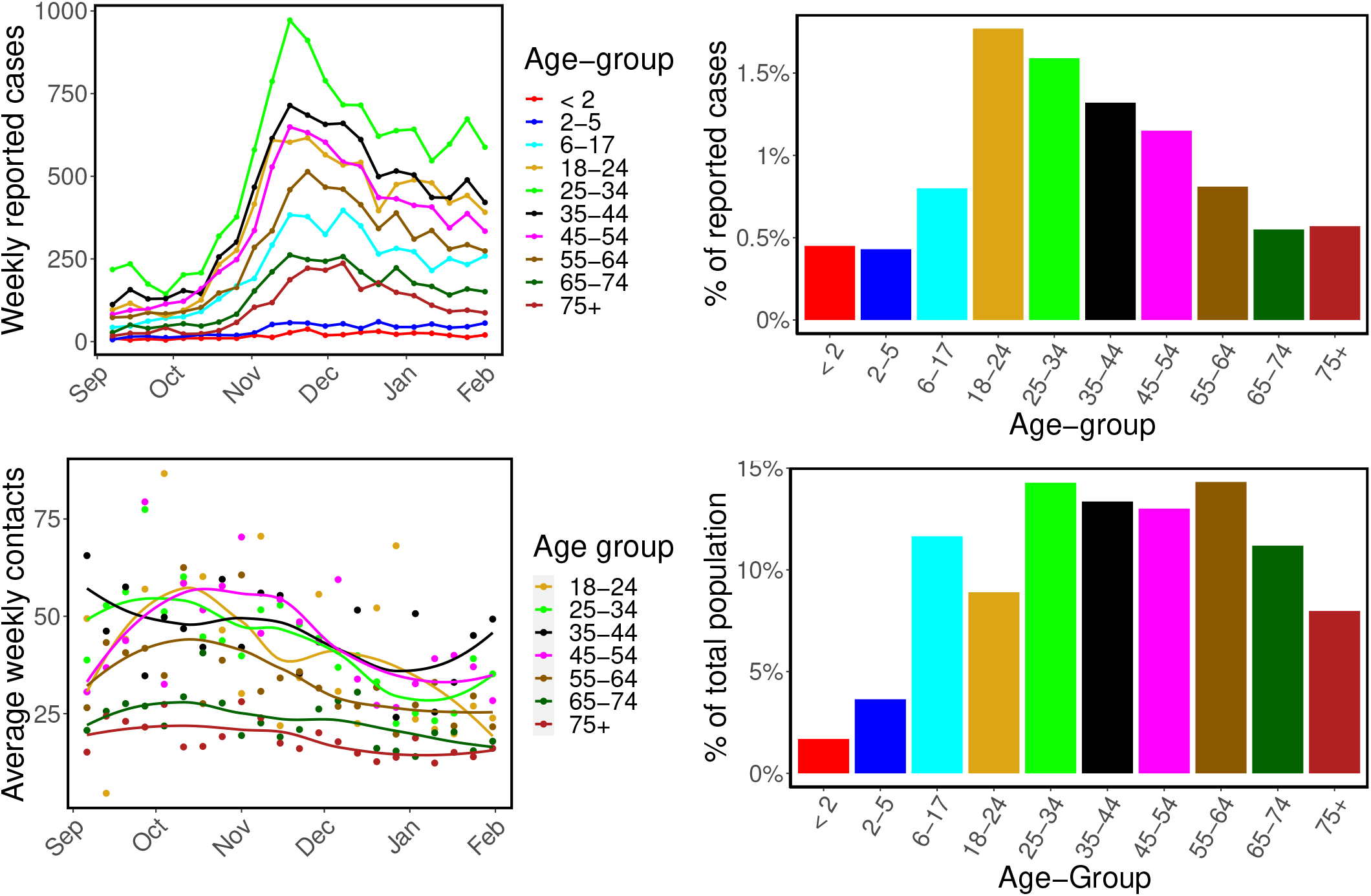
Reported cases of COVID-19 and contact rates in BC. Top left: Weekly reported cases of COVID-19 in BC by symptom onset date from September 2020 to January 2021. Top right: Cumulative reported cases by age group during study period, as a percentage of that age group’s population size. Bottom left: Average weekly contact rates by age group over time from September 2020 to January 2021. The dots are the computed average contact rates, while the solid lines are fits from local polynomial regressions with loess method (shown only for illustration of trends) [4]. Bottom right panel: BC population distribution by age group. The color codes are consistent across all the panels.

**Figure 3:**
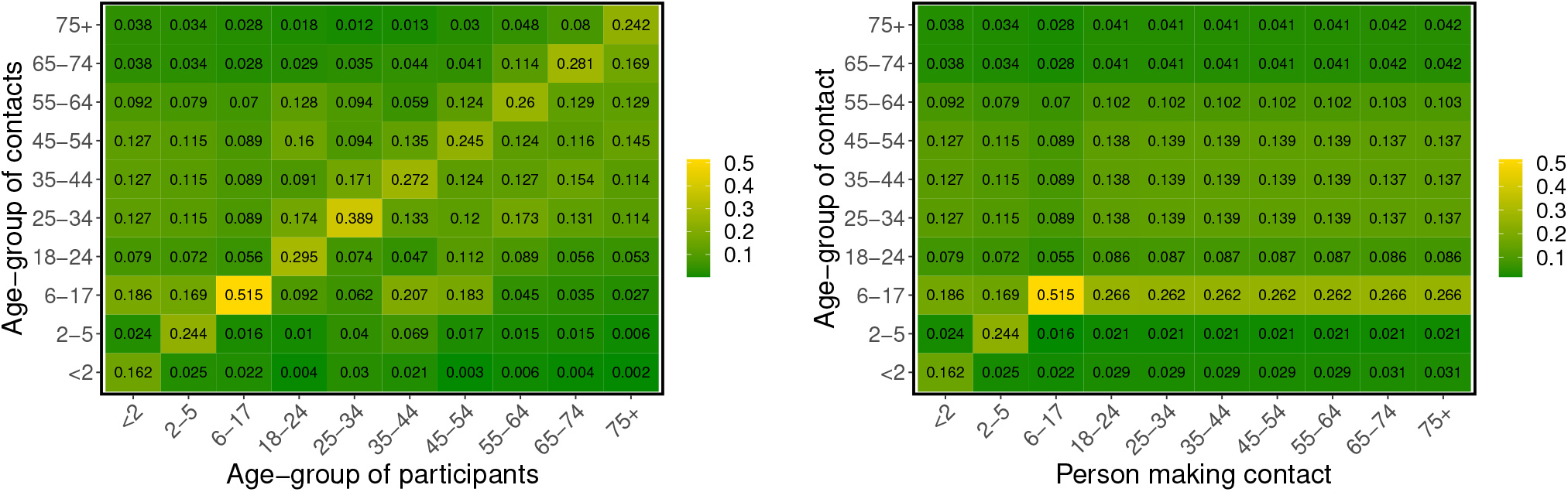
Population mixing patterns. *Heat maps showing the mixing patterns between individuals of different age groups as derived from the BC-Mix survey [1] (left panel) and influenza model of [16] (right panel). Each column represents the proportion of the total contacts of a specific age group made with each of the other age groups. Since the BC-Mix survey does not include individuals <* 18 *years, the columns for the age groups: <* 2 *years, 2-5 years, and 6-17 years in the BC-Mix matrix (left panel) were derived from the 2009 matrix (right panel)*.

Table 1 provides a description of the model parameters and their values. The population size of BC, stratified by the age groups of our model, is also given in Table 1. Since the case data considered in our fits start at September 2020, in order to coincide with the beginning of the BC Mix survey, rather than at the beginning of the COVID-19 epidemic in BC, the basic reproduction number *R*_0_ in the model (2.1) is the effective reproduction number *R*_0*t*_ in BC at the beginning of the period (September 2020 - January 2021). We compute the normalization constant *C*_*N*_ for the effective reproduction number *R*_0*t*_ using numerical bootstrapping based on the next-generation matrix approach of computing *R*_0_ [18, 60, 37]. To compute *C*_*N*_, we numerically construct the next-generation matrix for (2.1) with the transmission rate *β* as given in (2.2). Observe that (2.2) contains *R*_0_ and the parameter *C*_*N*_, which we want to compute. We set *R*_0_ = 1 and loop over a range of values of *C*_*N*_. For each value of *C*_*N*_, we compute the spectrum of the next-generation matrix and output the value of *C*_*N*_ for which the spectral radius of the matrix equals unity as our normalization constant. In other words, our desired normalization constant is the value of *C*_*N*_ that returns unity as the dominant eigenvalues of the next-generation matrix of the ODE system (2.1) when the basic reproduction number *R*_0_ = 1 in (2.2). It is important to mention that since the next-generation matrix depends on the contact rates and population mixing patterns, *C*_*N*_ also depends on these parameters. *C*_*N*_ incorporates the effect of population heterogeneity into the basic reproduction number and the transmission rate *β* (2.2). For all the results presented in this paper, individuals aged 0-17 are assumed to be 50% as susceptible as individuals 18 years or older [17, 7, 69]. Using a seropositivity rate of 4.5% at the start of the study period [15], we calculated the ascertainment fraction for reported cases to be 0.255 (i.e., 25.50% of all cases were assumed reported), which was then used when fitting to the age specific reported case numbers.

### 2.2 Data

The reported number of cases of COVID-19 in British Columbia were obtained from the British Columbia Centre for Disease Control (BCCDC) and stratified by the age groups of our model. Based on symptom onset date, we extracted these data from a line list generated by BCCDC Public Health Reporting Data Warehouse (PHRDW). Approximately 12.93% of the total reported cases during our study period do not have symptoms onset date. These cases were removed from our analysis. The collected data was incorporated into the model likelihood based on disease incidence (2.4). The case data used here covered the time period September 2020 to January 2021, inclusive. This time period was selected to match with the available contact rate data (see below), but also provides a wide range of COVID-19 dynamics for modeling.

We focused here on the dynamics of community transmission and therefore excluded residents of long-term care homes (LTCH). In particular, cases were removed if the transmission setting was specifically identified as a long-term care facility and the case was identified as a patient or resident (not staff). In the SEIR model (2.1), incidence is computed as the number of pre-symptomatic individuals (*E*_2_) transitioning to the infected class (*I*_1_). Figure 2 shows the weekly reported cases of COVID-19 in BC by age group from September 2020 to January 2021 (top left) and the percentage of cumulative reported cases for each age group within this period (top right). The 18-24 age group had the highest reported infection rate (by population size), with total reported cases during our study period corresponding to 1.77% of their population, followed by the 25-34 age group with 1.59%, and the 35-44 age group with 1.32% of their total population. The percentage of reported cases in the remaining age groups were; 45-54 years (1.15%), 55-64 years (0.81%), 6-17 years (0.80%), 75+ years (0.57%), 65-74 years (0.55%), *<* 2 (0.45%), and 2-5 (0.43%). We observe from this figure that the reported cumulative cases of COVID-19 was not uniform across age groups, possibly due to the variation in contact rates and mixing patterns between age groups, differences in reporting, and reduced susceptibility among younger individuals [17, 7, 69].

Heterogeneity in contact rates and mixing patterns among age groups are essential aspects of our model. These parameters are introduced through the average weekly contact rate vector *K* and the *N*_age_ *× N*_age_ mixing matrix *C*, which we compute from the BC-Mix survey [1]. This survey was initiated in September 2020 by the British Columbia Center for Disease Control (BCCDC) as part of ongoing public health surveillance for COVID-19. In the survey, BC residents 18 years of age and older were asked questions related to their daily contacts, response to non-pharmaceutical interventions such as the wearing of face masks and physical distancing, and vaccination. We computed the average weekly contact rate based on survey responses to the question: *“How many people did you have in-person contact with between 5 am yesterday and 5 am today?”*, where an *in-person contact* is defined as either having an in-person two-way conversation with three or more words or physical skin-to-skin contact, e.g. a handshake, hug, kiss, or contact sports (see [1] for more details). For each week from September 2020 to January 2021, contact rates were stratified by the age groups: 18-24 years, 25-34 years, 35-44 years, 45-54 years, 55-64 years, 65-75 years and 75+ years, and then used to compute the average contact made by each age group weekly. These results are presented in Figure 2 (bottom left), where the dots are the computed average contact rates and local polynomial regression lines using the *loess* method [4] are included to illustrate time trends. Since the BC-Mix survey is only available for individuals 18 years and above, the average weekly contact rates for the age groups *<* 2 years, 2-5 years, and 6-17 years were fixed at 5.5280, 12.4169, and 13.4486, respectively, throughout the epidemic period of September 2020 - January 2021. We derived these contact rates from the original influenza model of [16], which was based on empirical contact data from 2009 for the lower mainland of BC. Although it is difficult to determine if these contact rates remain fully representative in the current time period, our model structure allows flexibility (through scaling parameters, described below) in how contacts impact transmission. The bottom right panel of Figure 2 shows the BC population size distribution by age group [6] (see Table 1 for actual values).

We constructed the population mixing matrix *C* from the BC-Mix survey data. Respondents were asked to provide their own age, as well as the ages of their first ten contacts from the previous day. This information was used to compute, for each age group, the proportion of their total contacts that were made with each of the other age groups. This corresponds to the columns in the *N*_age_ *× N*_age_ population mixing matrix *C*, each of which sum to 1. Unlike the average contact rates by age group, which we computed weekly, only one mixing matrix was constructed from the survey data based on the total data from September 2020 to January 2021. In Figure 3, we present the population mixing matrix *C* derived from the BC-Mix survey (left panel) and, for comparison (and later use in modeling), the corresponding matrix from the 2009 influenza study of [16] (right panel). For convenience, we refer to the mixing matrix derived from the influenza model of [16] as the *2009 matrix*, and the one constructed from the BC-Mix survey as *BC-Mix matrix*. Note that in order to complete the first three columns of the BC-Mix matrix (i.e., for individuals under 18 years of age), we adapted data from the 2009 matrix for age groups *<* 2 years, 2-5 years, and 6-17 years. We observe a pattern of assortative mixing from the BC-Mix matrix (i.e., contact proportions are higher within age groups than between age groups), although this pattern is not as evident in the 2009 matrix. It is also apparent that children *≤* 5 years of age make a relatively large proportion of their contacts with adults of child-bearing age (25-54 years) and with adolescents 6-17 years of age. This pattern likely reflects family contacts among children, parents and siblings.

### 2.3 Bayesian inference

We fit our age-structured model (2.1) to the weekly reported cases of COVID-19 in BC (top left panel of Figure 2) for all the age groups simultaneously, using a Bayesian inference framework and the RStan package in R version 3.6.3 [55]. For each age group, the likelihood is constructed according to 

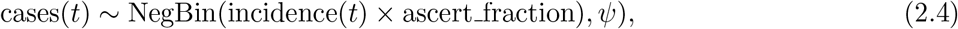

where NegBin(.) is the negative binomial distribution, cases(*t*) and incidence(*t*) are the weekly reported cases of COVID-19 in BC and the incidence computed from our SEIR model (2.1), respectively. The ascertainment fraction is given by ascert_fraction and *ψ* is the over-dispersion parameter. This framework allows us to incorporate our prior knowledge into the model parameters and gives us the ability to evaluate probabilistic statements of the data given the model.

To translate contact rates computed from the BC-Mix survey into impacts on transmission, we introduce scaling parameters for the average weekly contact rates, which are estimated for each age group from the fit. We also incorporate as a parameter the prevalence of COVID-19 in BC at the beginning of our study period (September 2020). As a validation step we simulated incidence data from our SEIR model (2.1) using known parameter values and tested our model’s ability to recover these values. The resulting posterior distributions were inspected for biases and coverage of the true parameters. All inferences performed in this paper were done using the Variational Bayes (VB) method with the *meanfield* algorithm implemented in RStan [56, 68], where the Bayesian model parameters were the initial total disease prevalence in the population and the scaling parameters for the contact rates. Other fixed parameters of the model are given in Table 1. The VB method was compared to the adaptive Hamiltonian Monte Carlo method No-U-Turn sampling and found to produce comparable estimates of the posterior distribution with significant reduction in total computation time [31]. Although, we set the maximum number of iterations for optimizing the evidence-lower bound (ELBO) in VB to 10,000, the mean and/or median ELBO usually converge in 1000 - 3000 iterations of the stochastic gradient ascent algorithm (see [32, 68, 28] for more information about ELBO in the variational Bayes method).

We considered four variations of the above Bayesian model based on the age-specific contact rates and the scaling parameters used to remove biases from the contact rates computed from the BC-Mix survey. These specific models, in order of increasing parameter flexibility, are:

1. A single scaling parameter for contact rates across all age groups and a fixed contact rate for each age group throughout the study period (SSFR).
2. Age-specific scaling parameters and a fixed contact rate for each age group (ASSFR).
3. A single scaling parameter across age groups and time-dependent average weekly contact rates for each age group (SSVR).
4. Age-specific scaling parameters and time-dependent average weekly contact rates (ASSVR).

Performing inference with each of these models provided a framework to understand the independent and combined effects of contact rates (fixed or time-varying) and scaling parameters (one for all age groups, or one for each age group) on the posterior predictive distribution and to determine which of the four models produces the best recreation of the observed data. We rank the models by comparing their leave-one-out predictions and standard errors, computed using the leave-one-out cross-validation (LOO) method [44, 21, 61]. Further comparisons were done using the widely applicable information criterion (WAIC) method [63, 20]. The fixed contact rates used for the SSFR and ASSFR models were given as the average of the weekly contact rates for each age group from September 2020 to January 2021. As an additional model to compare the impact of the contact mixing matrix on the posterior predictive distribution, we constructed each of the above four models using the 2009 survey-derived contact matrix as opposed to the BC-MIX survey-derived contact matrix (see Supplementary material A.3).

## 3 Results

We fit our age-structured SEIR model (2.1) to the weekly reported cases of COVID-19 in BC from September 2020 to January 2021, shown in the top left panel of Figure 2. Of primary interest was the model fits based on 2020/2021 contact rates from the BC-Mix survey [1]; however, we also compared fits based on previous contact network data collected during 2009 [16].

### 3.1 Model fitting scenarios

In Figure 4, we present our model fits for the BC-Mix matrix (left panel of Figure 3) for the four models described above for selected age groups (2-5 years, 35-44 years, and 55-64 years) and the total reported cases. The fits for the remaining age groups are presented in Supplementary material A.1. In the first scenario (SSFR), only a single scaling parameter is estimated across age groups (0.949, 90% CrI: 0.936 - 0.964; initial prevalence estimate = 9,827, 90% CrI: 8,455 - 11,245) and contact rates for each age group are fixed over time. This model predicts an approximately linear increase in cases over time, which fails to capture the true trends in the data. Similar results are shown for the ASSFR model, which had separate age-specific scaling parameters (see Figure 5 and Table S1 of Supplementary material A.1; initial prevalence estimate = 5,488, 90% CrI: 5,173 - 5,806), but still used fixed contact rates for each age group. This scenario again predicts a linear increase in cases, although, in contrast to the SSFR model, the credible intervals of the predictions are narrower and the predicted median incidence crudely approximates the overall trend in the underlying data. Nevertheless, the ASSFR model still fails to capture the dynamic time trends in the case data.

**Figure 4:**
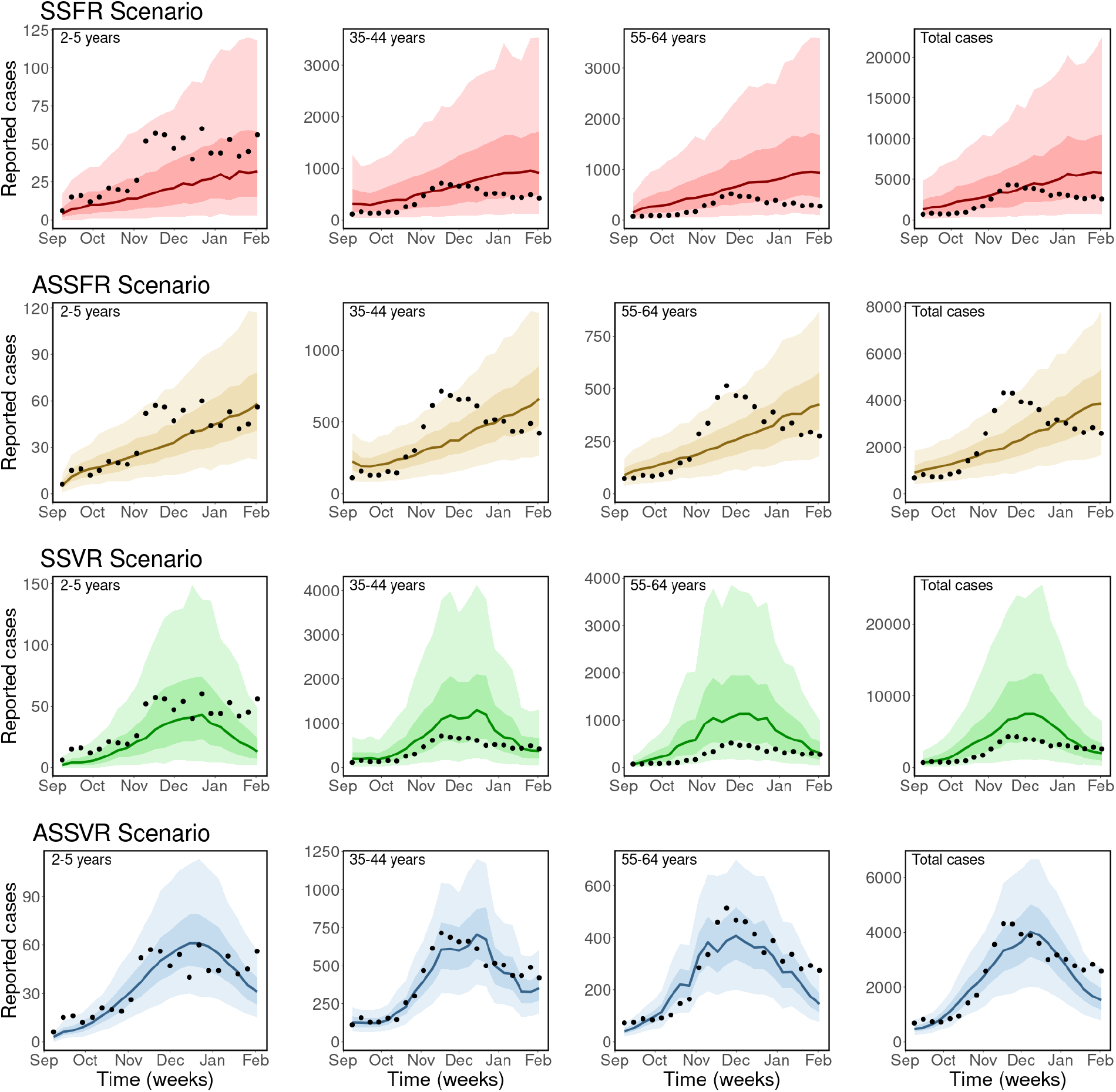
Observed and estimated case counts obtained using the BC-Mix matrix. *Top row (red): Single scaling parameter and fixed contact rates (SSFR). Second row (gold): Age-specific scaling parameter and fixed contact rates (ASSFR), third row (green): single scaling parameter and time-dependent contact rates (SSVR), and fourth row (blue): Age specific scaling parameter and time-dependent contact rates (ASSVR). The black dots are the weekly reported cases of COVID-19 in BC, the solid lines are the median predicted cases, the darker bands are the* 50% *CrI, while the lighter bands are the 90*% *CrI. Similar plots for the remaining age groups are provided in Supplementary material A*.*1*.

**Figure 5:**
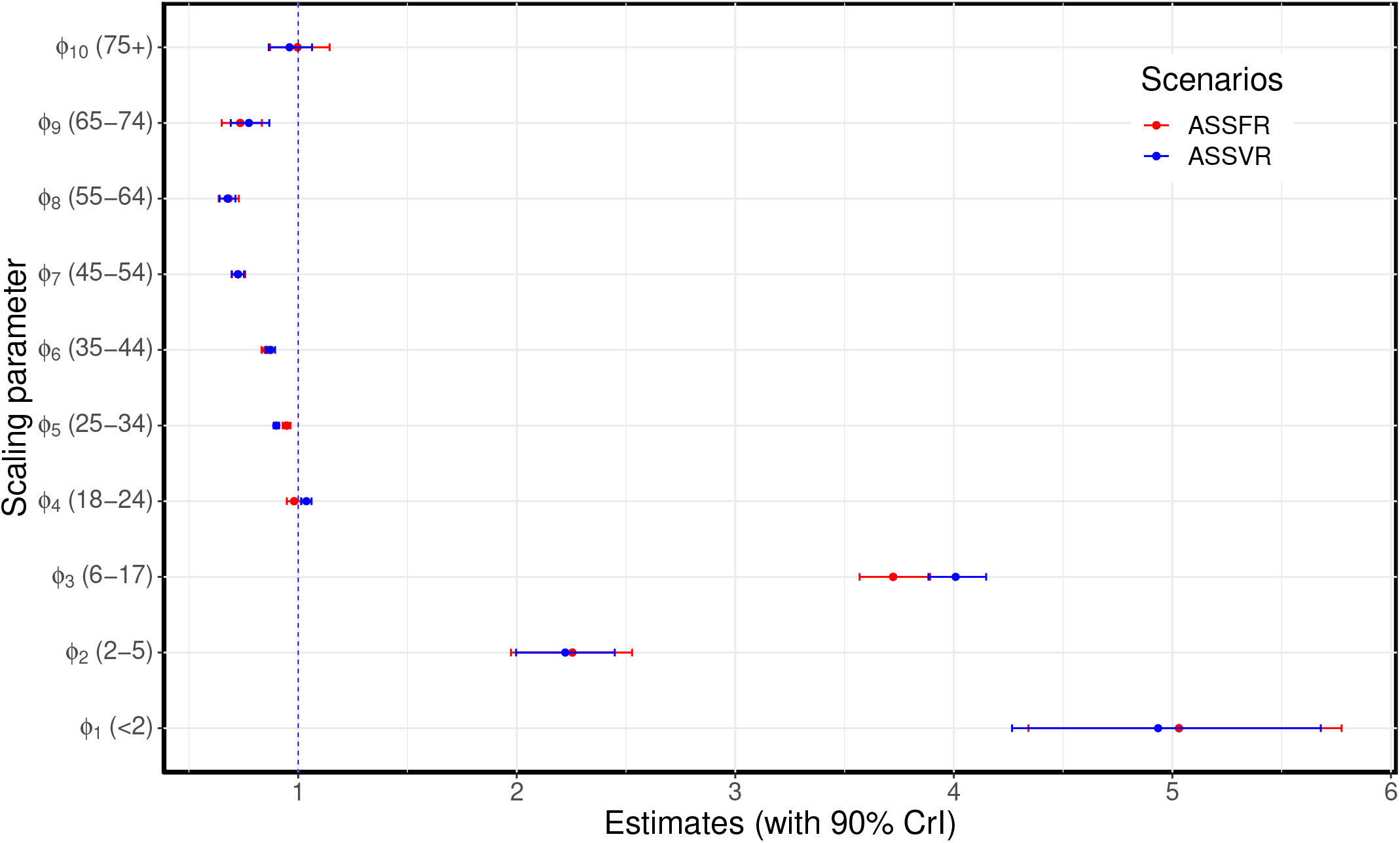
Estimated age-specific scaling parameters for the BC-Mix matrix. ASSFR model (red) and ASSVR model (blue). The blue vertical dashed line at 1 indicates where the scaling parameter has no effect on the fit. These estimates are presented in Table S1 of Supplementary material A.1.

The remaining results in Figure 4 show the SSVR and ASSVR scenarios, where time-dependent average weekly contact rates were used. In these scenarios, our model was able to capture the trends in the reported cases. Due to the single scaling parameter (0.930, 90% CrI: 0.919 - 0.940; initial prevalence estimate= 5,039, 90% CrI: 4,430 - 5,496), the SSVR model was unable to accurately predict case numbers, even though it captured the temporal trends in the data. It either overestimated or underestimated reported cases with large credible intervals. In contrast, the ASSVR model (initial prevalence estimate= 2,824, 90% CrI: 2,690 - 2,959; age-specific scaling parameters provided in Figure 5 and Table S1 of Supplementary material A.1), captured both the trends in reported cases and the case numbers with a higher accuracy compared to the other scenarios. In this scenario, we used age-specific scaling parameters together with time-dependent average weekly contact rates. Of all the four models considered, ASSVR with age-specific scaling parameters and time-dependent contact rates provided the best reconstruction of the reported cases data (fourth row of Figure 4). In addition, a comparison of the leave-one-out predictions and standard error for the four models, computed using the leave-one-out cross-validation (LOO) method [21, 61], ranked the ASSVR model as having the highest predictive accuracy, followed by ASSFR, SSVR, and then SSFR. A similar ranking was obtained using the widely applicable information criterion (WAIC) method [63, 20]. See Table S2 of Supplementary material A.1 for detailed output of the model comparison. Taken together, these results suggest that the time-dependent weekly contact rates are required to accurately capture the trends in the reported cases, while the age-specific scaling parameters allow the fits to properly match actual numbers of reported cases. Similar results were obtained using the 2009 mixing matrix (right panel of Figure 3). These results are presented in Supplementary material A.3.

### 3.2 Scaling contact rates

Comparisons of the estimated age-specific scaling parameters for the ASSFR and ASSVR models based on the BC-Mix contact data are shown in Figure 5 (similar results are shown for the 2009 matrix in Figure S10 of Appendix A.3). The blue vertical dashed line at 1 indicates where the scaling parameter has no effect on the fit. When the scaling parameter is less than 1, it indicates a reduction in the contribution the observed contact rates have to transmission; conversely, scaling parameters greater than one indicate an increase in the contribution observed contact rates make to transmission. We observe that the estimated scaling parameters for the ASSFR and ASSVR models are generally similar and closer to 1 for all age groups except *<* 2 years, 2-5 years, and 6-17 years. Overall, relatively little re-scaling was needed for contact rates computed from the BC-Mix survey, suggesting that the survey was able to capture contact activity relevant for COVID-19 transmission. However, the estimated scaling parameters for ages *<* 2 years, 2-5 years, and 6-17 years were notably greater than 1 and with wider 90% credible intervals. This suggests that self-reported contact rates for youngest age groups tended to under-estimates the level of contact relevant to COVID-19 transmission.

Recall that these youngest age groups were not available in the BC-Mix survey and that we used instead the contact rates derived from the original influenza model of [16] for age 0-17 years. It is possible that these contact rates, recorded during 2009, were not fully representative of contacts made during our 2020/2021 study period. Nevertheless, model fits for the 0-17 ages groups were generally close to the observed data S4, suggesting the scaling parameters were able to largely accommodate any discrepancies. We also assumed in our model that individuals in these youngest age groups are only 50% as susceptible as adults. This may contribute to the high estimates in the scaling parameters as reduction in susceptibility would lead to fewer infections, and the scaling parameters may reflect a correction for this. The estimates of the age-specific scaling parameters shown in Figure 5 are presented in Table S1 of Supplementary material A.1. Similar results are presented in Tables S3 of Supplementary material A.3 for the 2009 matrix.

### 3.3 Model validation and projection

Next, we generated short-term projections of new cases from our model fits to help validate our modeling framework. We fit our model to a subset of the reported cases (September to December 2020), and projected forward to predict cases for the month of January 2021 (4 weeks). We then compared the predicted cases against actual reported cases for this 4-week period. The ASSVR model was used for this study with the results presented in Figure 6 for all the age groups and total reported cases.

**Figure 6:**
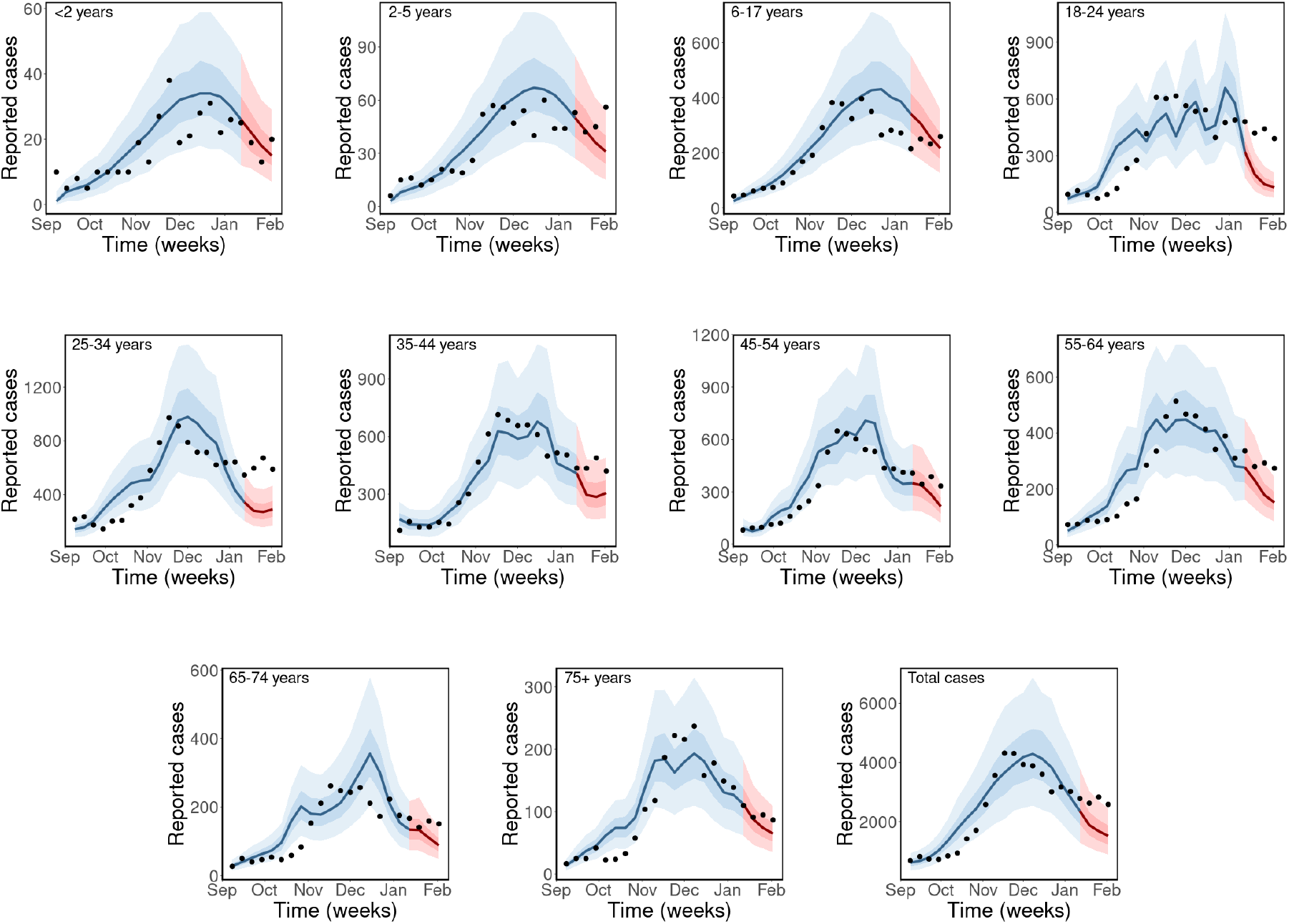
Model validation and projection. *Model fits for the period from September to December 2020 (in blue) and projection for January 2021 (in red) with* 50% *and* 90% *credible interval (CrI) bands, using the BC-Mix matrix. The solid lines are the median predicted cases, narrow bands are* 50% *CrI, and wider bands are* 90% *CrI. Black dot are weekly reported cases of COVID-19 in BC*.

Our model was able to accurately predict trends in most, but not all, age groups. True case numbers fell within the 90% credible intervals of our predictions for most age groups, except 18-24 and 25-34 years, where our model slightly underestimated reported cases. It is important to mention that for the prediction period January 2021, we used the average weekly contact rates for this month as computed from the BC-Mix survey. In reality, contact rates for future dates would not be available and would need to be hypothesized based on historical contact rates for different possible scenarios.

## 4 Discussion

We developed an SEIR-type age-structured model that explicitly incorporates contact rate data and mixing patterns computed from the BC-Mix survey [1] for studying the transmission dynamics of COVID-19. Our model builds upon a framework that used age- and activity-structure to study the dynamics of the 2009 H1N1 virus in the Greater Vancouver area of British Columbia, Canada [16]. It divides the population of BC into: susceptible (*S*), exposed (not infectious *E*_1_ and pre-symptomatic infectious *E*_2_), infected (*I*_1_ and *I*_2_) and recovered compartment (*R*), and stratifies each into ten age groups: *<* 2 years, 2-5 years, 6-17 years, 18-24 years, 25-34 years, 35-44 years, 45-54 years, 55-64 years, 65-74 years, and 75+ years. Unlike the other age-structured models [46, 47, 13, 12, 11] used to study the dynamics of COVID-19, where the contact rates are fixed throughout the epidemic period, our model uses time-varying weekly contact rates computed directly from ongoing survey data (BC Mix COVID-19 survey (BC-Mix) [1]). Hence, the effects of non-pharmaceutical interventions (NPIs) on the activity level of each age group are implicitly captured by the contact rates computed from the survey. Further, we use a population mixing matrix, derived from the same survey data, which specifies the proportion of total contacts of an age group that is made with each of the other age groups. For simplicity, we elected to use a single mixing matrix for the entire study period from September 2020 to January 2021, which assumes population mixing patterns in BC were roughly constant. For comparison, we also implemented our model with the age mixing matrix of [16] used to study 2009 H1N1 transmission in the Greater Vancouver area. It is likely that patterns of mixing among age groups change over time (e.g., due to age-specific seasonal behaviours, responses to public health measures, etc.); an interesting extension of this work would be to explore time trends in the mixing matrix itself and assess how this impacts the observed transmission dynamics.

Using a Bayesian inference framework, we calibrate our model to weekly reported cases of COVID-19 in BC from September 2020 to January 2021. We show that our framework is robust to population mixing patterns, with similar results found using the current (2020/2021 BC-Mix survey) and previous (2009 influenza mixing survey) mixing matrices. Our use of scaling parameters to translate self-reported contact rates in each age group into impacts on transmission helps to account for aspects of self-reported survey data, such as recall bias and challenges in defining which contacts actually lead to transmission. We considered four different Bayesian models based on the scaling parameters (single scaling parameter vs age-specific scaling) and on the contact rates themselves (fixed contact rates vs time-varying rate). Fixed contact rates for each age group, predicted an approximately linear growth in cases and failed to capture observed trends in the data. With time-varying contact rates, our model captured trends in reported cases, but significantly under- or over-estimated the actual values. Only the combination of age-specific scaling parameters and time-dependent contact rates provided adequate fits to observed cases data. In addition, the Bayesian model with age-specific scaling parameters and time-varying average weekly contact rates was ranked the most preferred using the leave-one-out cross-validation (LOO) and the widely applicable information criterion (WAIC) methods. These results show that, beyond incorporating population heterogeneity from contact rates and mixing patterns, it is also essential to use time-varying contact rates and allow age-specific scaling of these rates.

We validated our modeling framework by fitting our model to the cases data from September to December 2020 and projected forward to predict the reported cases for January 2021 using the posterior distribution of the estimated parameters. Our projections provide moderate to good fits to the observed reported cases. In most age groups, we captured the reported cases within the 90% credible intervals of our model prediction. As expected, our projections are highly dependent on the underlying contact rates; as a result, we were able to capture the true trends in cases particularly for older age groups, which are well represented in the contact survey data. In contrast, the lower response rates among younger adults (18-34 years) in the BC-Mix survey may have resulted in contact rates (and hence, projections) that were less representative of the true trends. A related limitation of our framework is that it requires one to specify the future weekly contact rates in order to project future cases. One way to approach this issue would be to use contact rates from previous weeks and assume these will be representative of future contact rates. Such projections would be interpreted as a possible scenario if future contact rates remained similar to historical levels. A distinct benefit of our model is that future projections can be made based on clear assumptions about contact rates in the underlying population, as opposed to more challenging assumptions about changes in transmission rates (which could be due to many factors).

Our modeling framework is based on deterministic ODE equations, which are well suited to capturing smooth changes in transmission but may not be suitable for sudden fluctuations. The sensitivity to underlying contact rates can be a limitation if contact data are, for any reason (including poor data quality) unstable. Fluctuating contact rates could lead to unrealistic predictions as the model tries to capture these dynamics with a series of small exponential growths and decays. This appears to be the case for our 18-24 age group, which typically had unstable contact rates due to small numbers of survey respondents of this age.

We show the importance of including population-based contact rates in modeling of COVID-19. In this study, we used self-reported rates of close contact from the BC-Mix survey which began during September 2020 and is (as of October 2021) still ongoing. By including such contact information, it removes the need to otherwise estimate how contact rates might change over time due to, for example, non-pharmaceutical interventions (NPIs), such as restrictions on gathering sizes, travel, and closure of businesses (e.g., stores, restaurants, etc.). Yet, this approach is not without limitations. Certain NPIs, such as face masks and handwashing, may reduce transmission without affecting contact rates. It is possible to include additional parameters in our model to account for changes in the probability of infection per contact, and this would be an interesting area for further work. In this paper, we take a more general approach and introduce scaling parameters that ‘translate’ self-reported contact rates into contributions to transmission, by age group to account for possible biases in the survey data. Another limitation to note is that current contact rates for the age groups *<* 2 years, 2-5 years and 6-17 years were not available (BC-Mix survey respondents must be 18+ years of age) and so were instead derived from the previous influenza model of [16]. These contact rates were computed during 2009 and thus may no longer represent the true pattern of population mixing in BC. Although the scaling parameters for these age groups seems to largely account for any discrepancies, it would be preferable to have current contact rates for all age groups in our model.

Our modeling framework focuses on age-related patterns of contact and transmission and does not consider other possible sources of variation. For example, we used average provincial contact rates for each age group and did not explore regional differences or variation by socio-economic or occupation group (e.g., essential workers). We are currently developing a variation of this SEIR model that focuses on the meta-population dynamics of transmission within and among health regions of BC. Another interesting extension of this work would be to use time-dependent scaling parameters and mixing matrices. By allowing these parameters and matrices to vary, the model may be better able to capture changes in mixing patterns among age groups and how contact rates translate into impacts on transmission. The impact of certain non-pharmaceutical interventions may vary among age groups, particularly for measures such as closure of schools and businesses; time-varying mixing and scaling would reduce the need to adjust the model fits specifically to reflect the implementation of these measures. Other interesting directions, already under investigation by our team, include incorporating the effect of COVID-19 vaccination and variants of concern (VoCs) in thaais modeling framework. Certain variants have been shown to be more contagious than the original SAR-CoV-2 virus and responsible for spikes in reported cases during spring/summer of 2021 in many parts of the world. Yet, at the same time, vaccination has substantially reduced transmission in many populations. These factors had negligible impact on the results presented here, given that our data ended January 2021, but are of critical importance when looking at later time periods. Indeed, the effects of VoCs and vaccination may be highly age specific and our modeling framework is thus naturally suited to incorporating these elements.

## 5 Conclusion

Incorporating real-time contact survey data into models of COVID-19 dynamics has the potential to provide more rapid and improved estimates of infection rates and projected trends. Our study shows how empirical contact rate data can be integrated into a transmission modeling framework and used to capture age-specific trends in cases. Further work could consider how different types and levels of contact, as reported through survey data, contribute to the transmission dynamics of COVID-19.

## Data Availability

All data produced in the present work are contained in the manuscript.

## Acknowledgement

S.I. thanks Sean Anderson and Julien Riou for their advice during the development of the Bayesian inference framework.

## A Supplementary material

### A.1 Model fits for the BC-Mix matrix

We present the Bayesian fits of the SEIR model (2.1) to the weekly reported cases of COVID-19 in BC from September 2020 to January 2021 (top left panel of Figure 2) for each age group and total reported cases. These results were obtained using the BC-Mix matrix (left panel of Figure 3).

**Figure S1:**
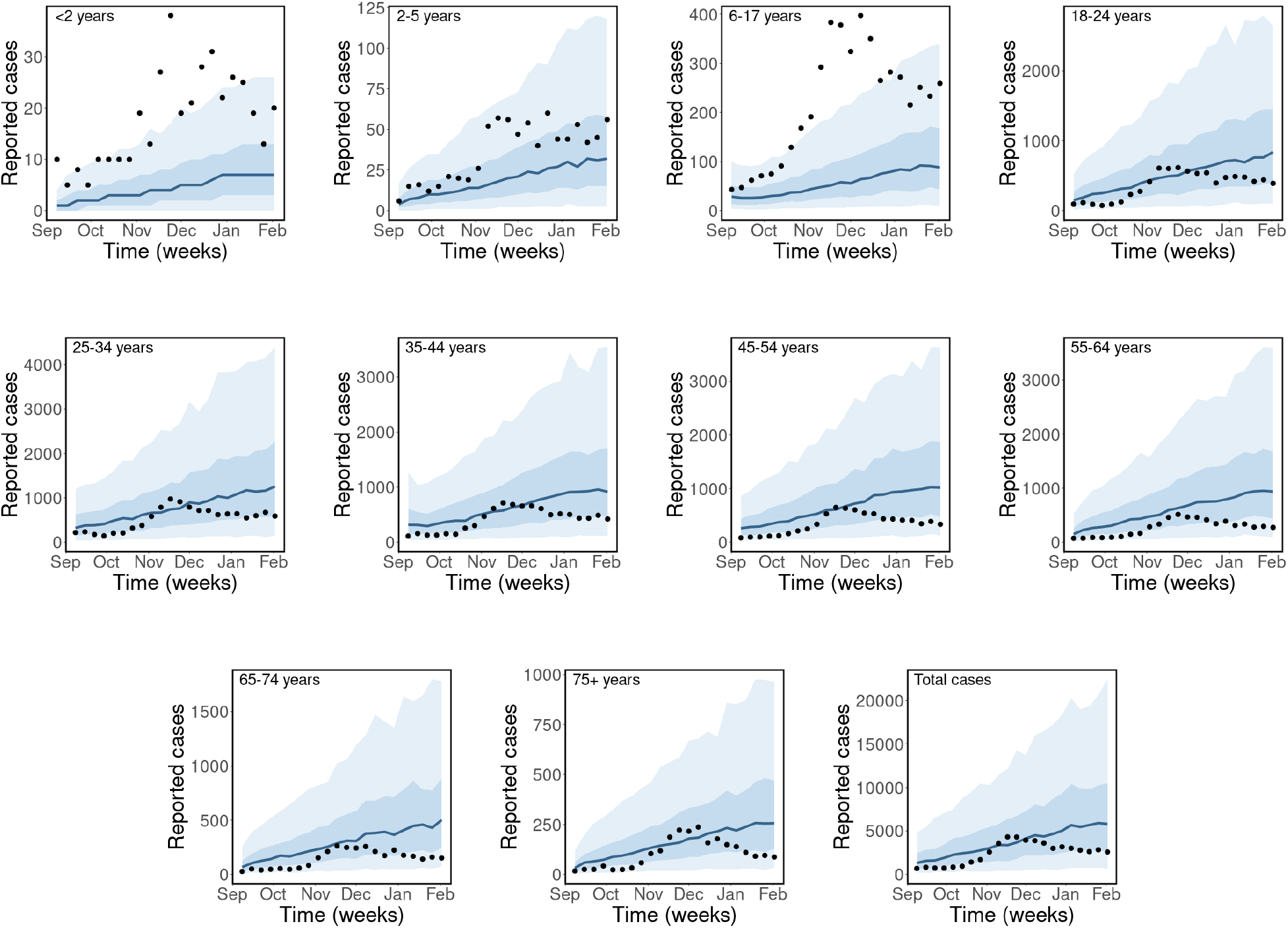
*Observed and estimated case counts by age group for SSFR model (single scaling parameter and fixed contact rate) for the BC-Mix matrix. The black dots are the weekly reported cases of COVID-19 in BC, the blue solid lines are the median predicted cases, the narrower bands are the* 50% *CrI, while the wider bands are the 90*% *CrI*.

**Figure S2:**
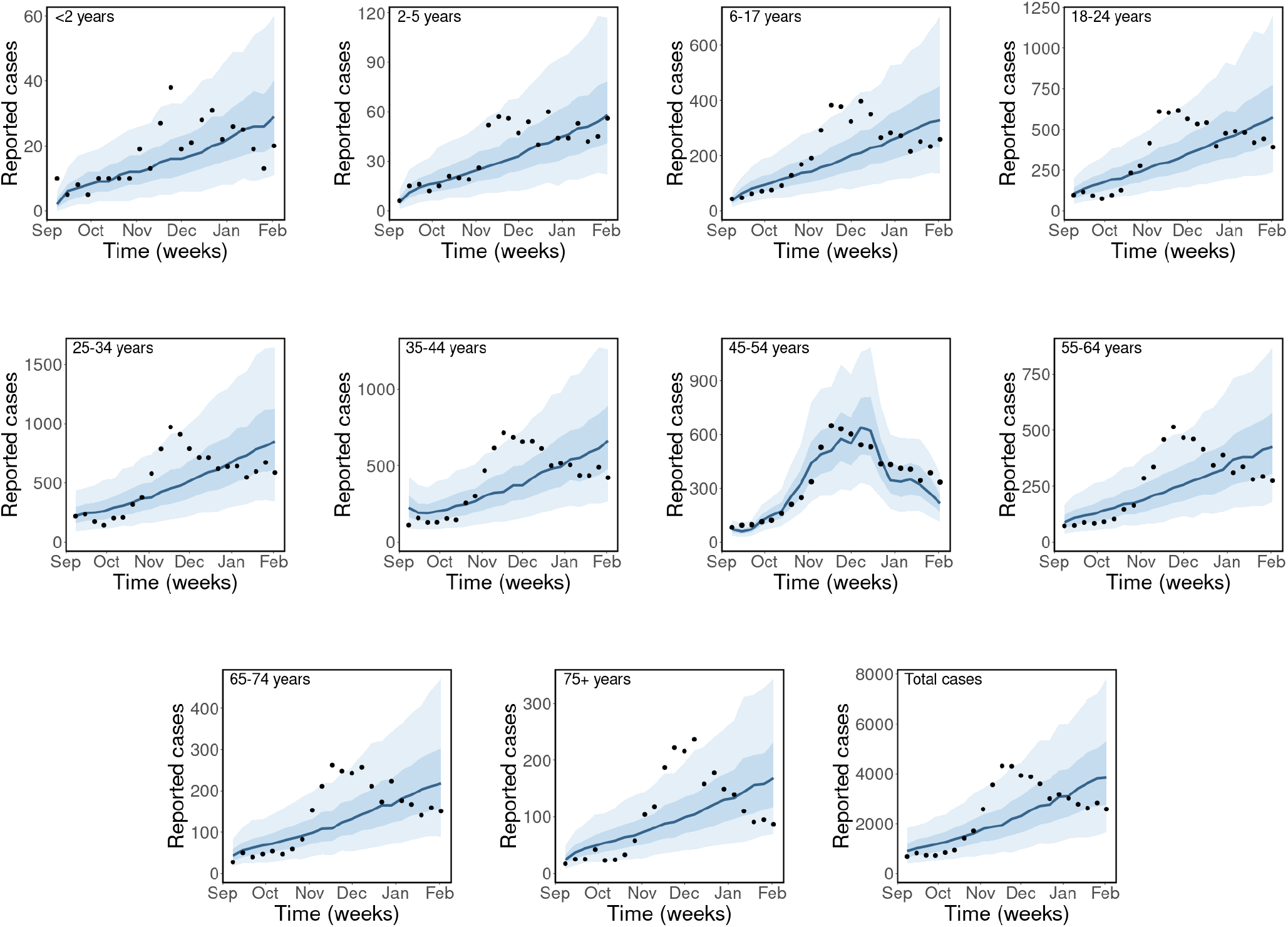
*Observed and estimated case counts by age group for ASSFR model (age-specific scaling parameters and fixed contact rates) for the BC-Mix matrix. The black dots are the weekly reported cases of COVID-19 in BC, the blue solid lines are the median predicted cases, the narrower bands are the* 50% *CrI, while the wider bands are the 90*% *CrI*.

**Figure S3:**
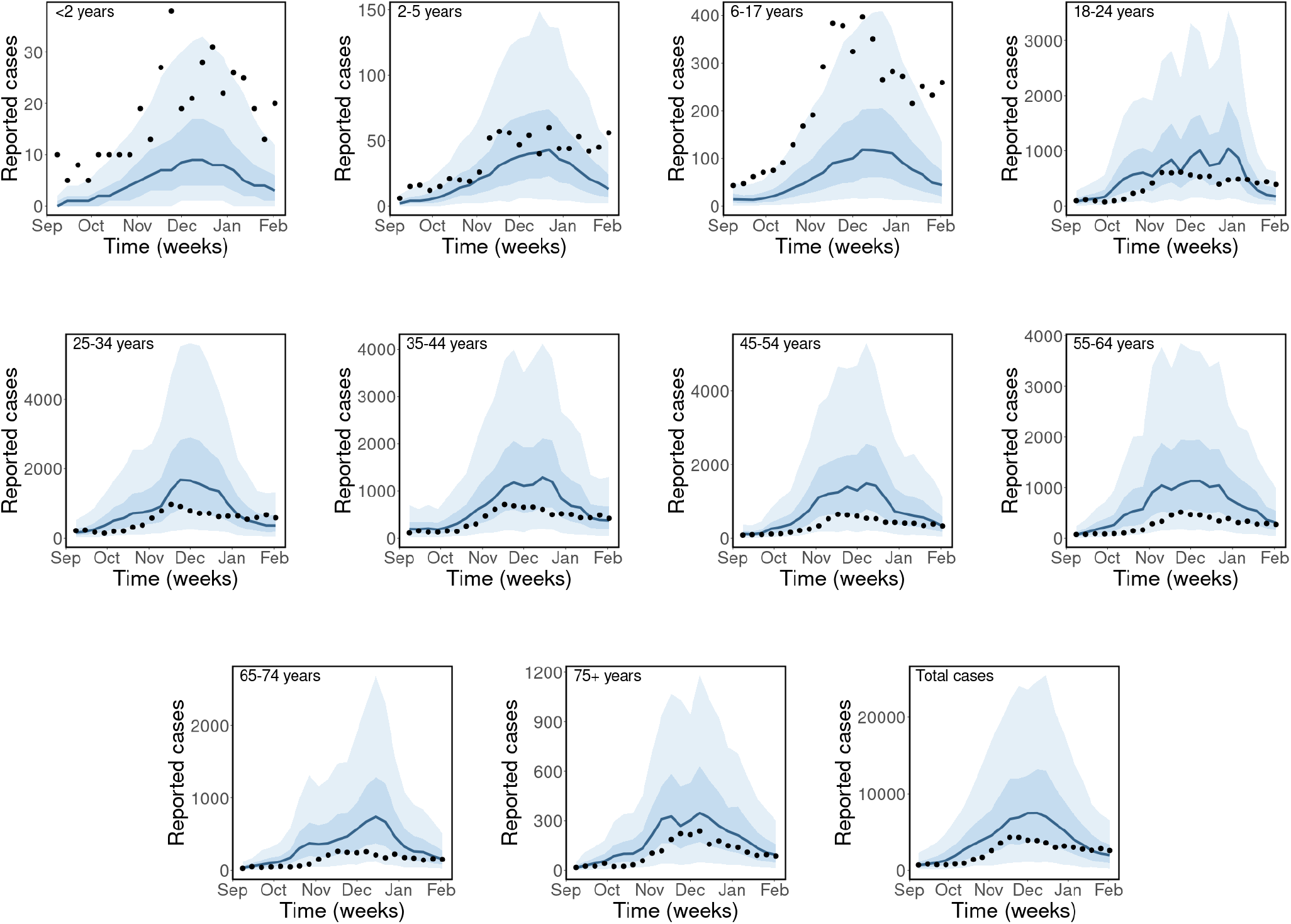
*Observed and estimated case counts by age group for the SSVR model (single scaling parameter and time-dependent average weekly contact rates) for the BC-Mix matrix. The black dots are the weekly reported cases of COVID-19 in BC, the blue solid lines are the median predicted cases, the narrower bands are the* 50% *CrI, while the wider bands are the 90*% *CrI*.

**Figure S4:**
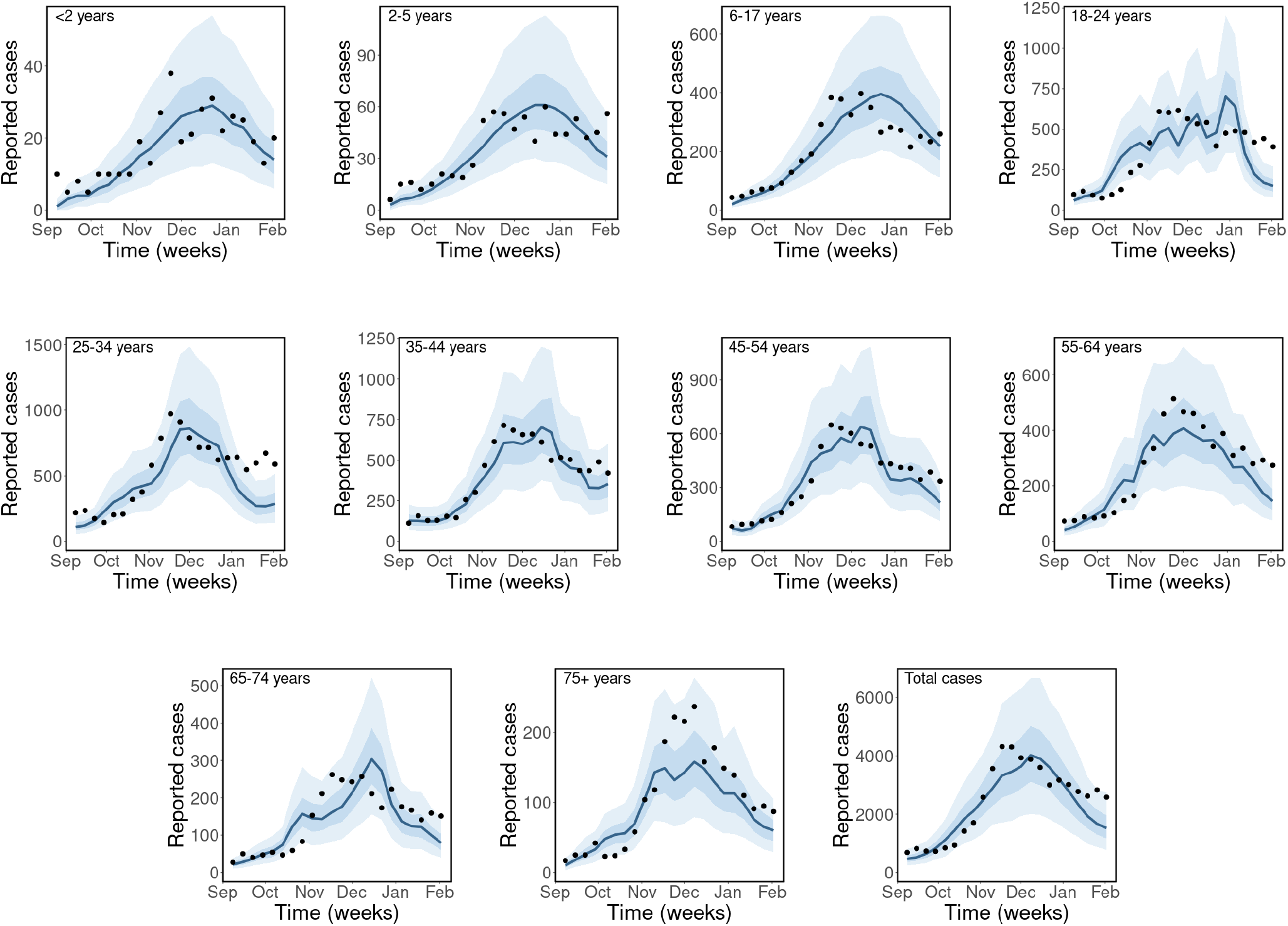
*Observed and estimated case counts by age group for ASSVR model (age-specific scaling parameters and time-dependent average weekly contact rates). The black dots are the weekly reported cases of COVID-19 in BC, the blue solid lines are the median predicted cases, the narrower bands are the* 50% *CrI, while the wider bands are the 90*% *CrI*.

**Table S1:** Estimated age-specific scaling parameters for the ASSFR and ASSVR models using the BC-Mix matrix.

| Scaling parameter | Age group | Estimates for ASSFR<br>scenario (90% CrI) | Estimates for ASSVR<br>scenario (90% CrI) |
| --- | --- | --- | --- |
| $\phi_1$ | < 2 years | 5.03 (4.34, 5.77) | 4.93 (4.27, 5.68) |
| $\phi_2$ | 2-5 years | 2.25 (1.97, 2.52) | 2.22 (2.00, 2.45) |
| $\phi_3$ | 6-17 years | 3.72 (3.57, 3.89) | 4.01 (3.88, 4.15) |
| $\phi_4$ | 18-24 years | 0.98 (0.95, 1.01) | 1.04 (1.01, 1.06) |
| $\phi_5$ | 25-34 years | 0.95 (0.93, 0.96) | 0.90 (0.89, 0.91) |
| $\phi_6$ | 35-44 years | 0.86 (0.83, 0.88) | 0.87 (0.85, 0.89) |
| $\phi_7$ | 45-54 years | 0.72 (0.70, 0.76) | 0.72 (0.70, 0.75) |
| $\phi_8$ | 55-64 years | 0.68 (0.64, 0.73) | 0.68 (0.64, 0.71) |
| $\phi_9$ | 65-74 years | 0.73 (0.65, 0.83) | 0.77 (0.69, 0.87) |
| $\phi_{10}$ | 75+ years | 1.00 (0.87, 1.14) | 0.96 (0.87, 1.06) |

### A.2 Model comparison

Here, we compare the estimated expected leave-one-out predictions and their standard errors, for the four models (SSFR, ASSFR, SSVR, and ASSVR), computed using the leave-one-out cross-validation (LOO) method [44, 21, 61] and the widely applicable information criterion (WAIC) method [63, 20]. A summary of the comparisons is shown in Table S2, where the ASSVR model, with age-specific scaling parameters and time-dependent average weekly contact rates, is ranked as the most preferred model. Followed by the ASSFR model (with age-specific scaling parameters and a fixed contact rate for each age group), then the SSVR model (with a single scaling parameter and time-dependent average weekly contact rates), and lastly the SSFR model with a single scaling parameter and a fixed contact rate for each age group (See the first column of Table S2).

**Table S2:** Summary of model comparison using leave-one-out cross-validation (LOO) and the widely applicable or Watanabe-Akaike information criterion (WAIC). Model ranking (in descending order) is shown in the first column. The difference between the expected log pointwise predictive density (elpd) and its standard error (se_elpd) for the best model and those of the remaining models are shown in the second column. In the third column, we have the Bayesian leave-one-out estimate of out-of-sample predictive fit (elpd_loo) and its standard error (se_elpd_loo). Lastly, the computed Watanabe-Akaike information criterion (waic) for each model is shown in the fourth column.

| Model | elpd_diff (se_diff) | elpd_loo (se_elpd_loo) | looic (se_looic) | waic |
| --- | --- | --- | --- | --- |
| ASSVR | 0.0 (0.0) | −1194.9 (20.0) | 2389.9 (40.0) | 2389.69 |
| ASSFR | −34.0 (14.7) | −1229.0 (20.8) | 2457.9 (41.6) | 2457.35 |
| SSVR | −200.5 (11.4) | −1395.4 (19.3) | 2790.8 (38.7) | 2790.77 |
| SSFR | −207.0 (14.2) | −1401.9 (19.4) | 2803.9 (38.8) | 2803.72 |

### A.3 Model fits for the 2009 matrix

Next, we present our Bayesian fit of the age-structured model (2.1) to the weekly reported cases of COVID-19 in BC from September 2020 to January 2021 (top left panel of Figure 2) for each age group and for total reported cases. These results were computed using the 2009 matrix (right panel of Figure 3) derived from the influenza model of [16].

Similar to the results obtained using the BC-Mix matrix, presented in Figure 4, we present the results for the 2009 matrix (right panel of Figure 3) for selected age groups and total reported cases. Overall the trends in the posterior distribution for the four models obtained using the 2009 matrix are similar to those of the BC-Mix matrix (Figure 4), although the estimated scaling parameters are slightly different. For the SSFR model, shown in the first row of Figure S5 (in red), the estimated initial prevalence is 8,315 (90% CrI: 7,309 - 9,533) and the estimated scaling fraction is 0.987 (90% CrI: 0.975 - 1.00). The results for the ASSFR model (age-specific scaling parameters and fixed contact rates) are shown in the second row of Figure S5, with the initial prevalence estimated as 6,095 (90% CrI: 5,815 - 6,363). The estimated age-specific scaling parameters are given in Figure S10 and Table S3. In the third row of Figure S5, we have the posterior predictive distribution for the SSVR model with initial prevalence estimated as 4,514 (90% CrI: 3,874 - 5,133) and the scaling parameter estimated as 0.956 (90% CrI: 0.943 - 0.967). Lastly, on the fourth row of Figure S5, we present the results for the ASSVR model (in blue), where age-specific scaling parameters are used together with time-dependent weekly contact rates. For this model, the estimated initial prevalence is 3,378 (90% CrI: 3,268 - 3,497). The estimated age-specific scaling parameters are given in Figure S10 and Table S3. Similar to the results for the BC-Mix matrix (Figure 4), the trends in the reported cases data are best reconstructed when age-specific scaling parameters are used together with time-dependent weekly contact rates, that is, the ASSVR model.

**Figure S5:**
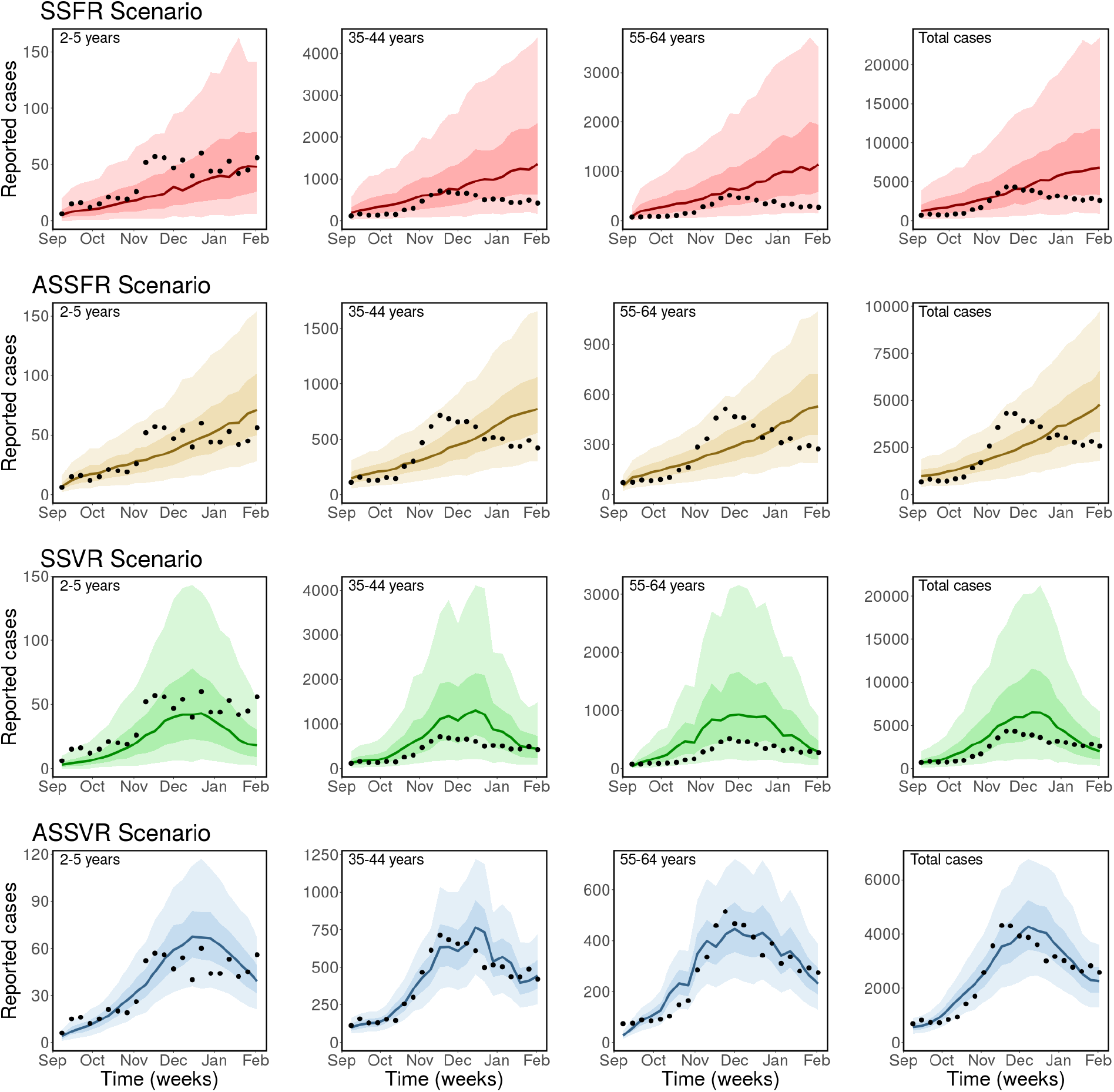
Observed and estimated case counts obtained using the 2009 matrix. *Top row (red): Single scaling parameter and fixed contact rates (SSFR). Second row (gold): Age-specific scaling parameter and fixed contact rates (ASSFR), third row (green): single scaling parameter and time-dependent contact rates (SSVR), and fourth row (blue): Age-specific scaling parameter and time-dependent contact rates (ASSVR). The black dots are the weekly reported cases of COVID-19 in BC, the solid lines are the median predicted cases, the darker bands are the* 50% *CrI, while the lighter bands are the 90*% *CrI*.

Next, we present the posterior predictive checks for the remaining age groups for the 2009 matrix.

**Figure S6:**
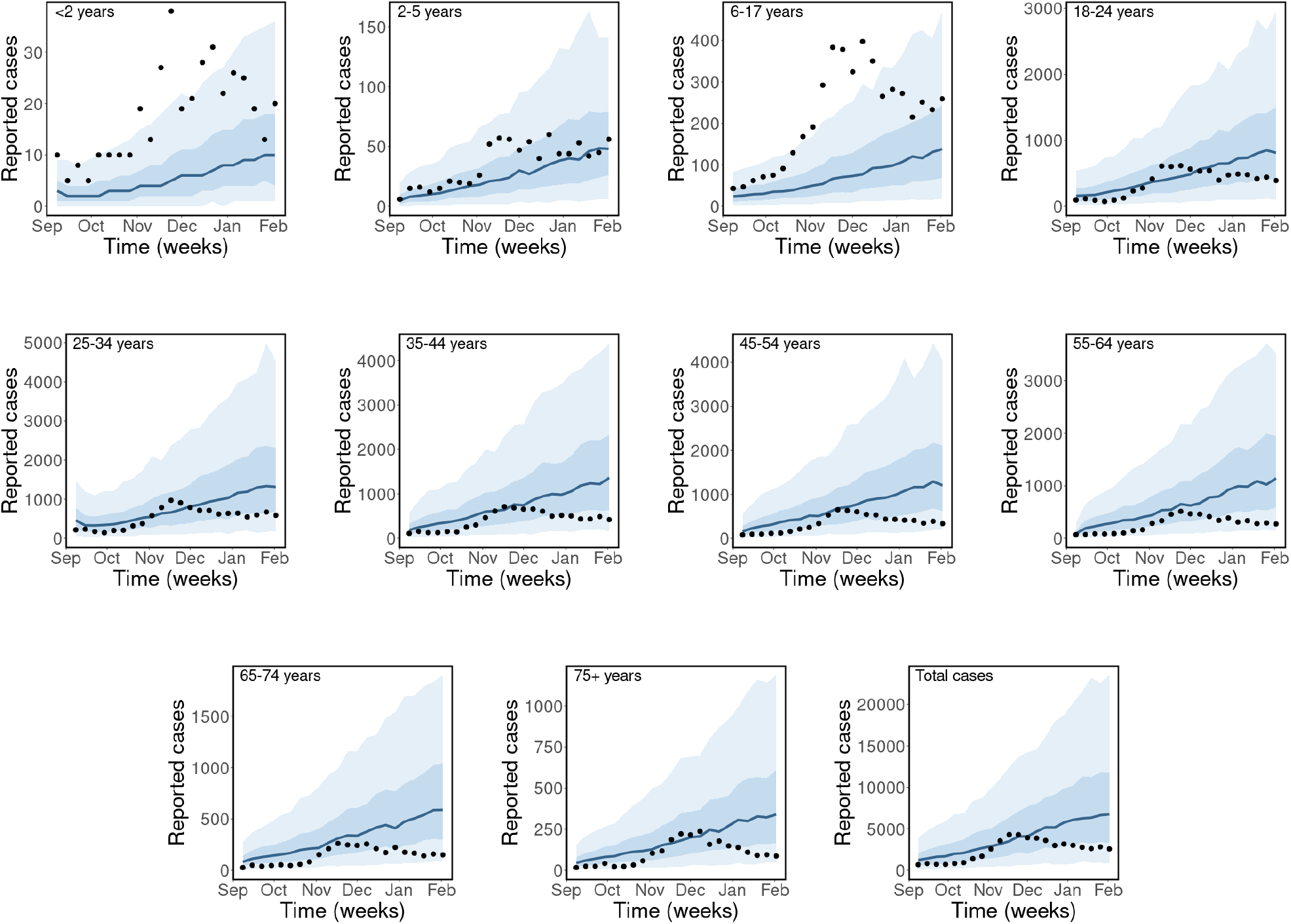
*Observed and estimated case counts by age group for the SSFR model (single scaling parameter and fixed average weekly contact rates) for the 2009 matrix. The black dots are the weekly reported cases of COVID-19 in BC, the blue solid lines are the median predicted cases, the narrower bands are the* 50% *CrI, while the wider bands are the 90*% *CrI*.

**Figure S7:**
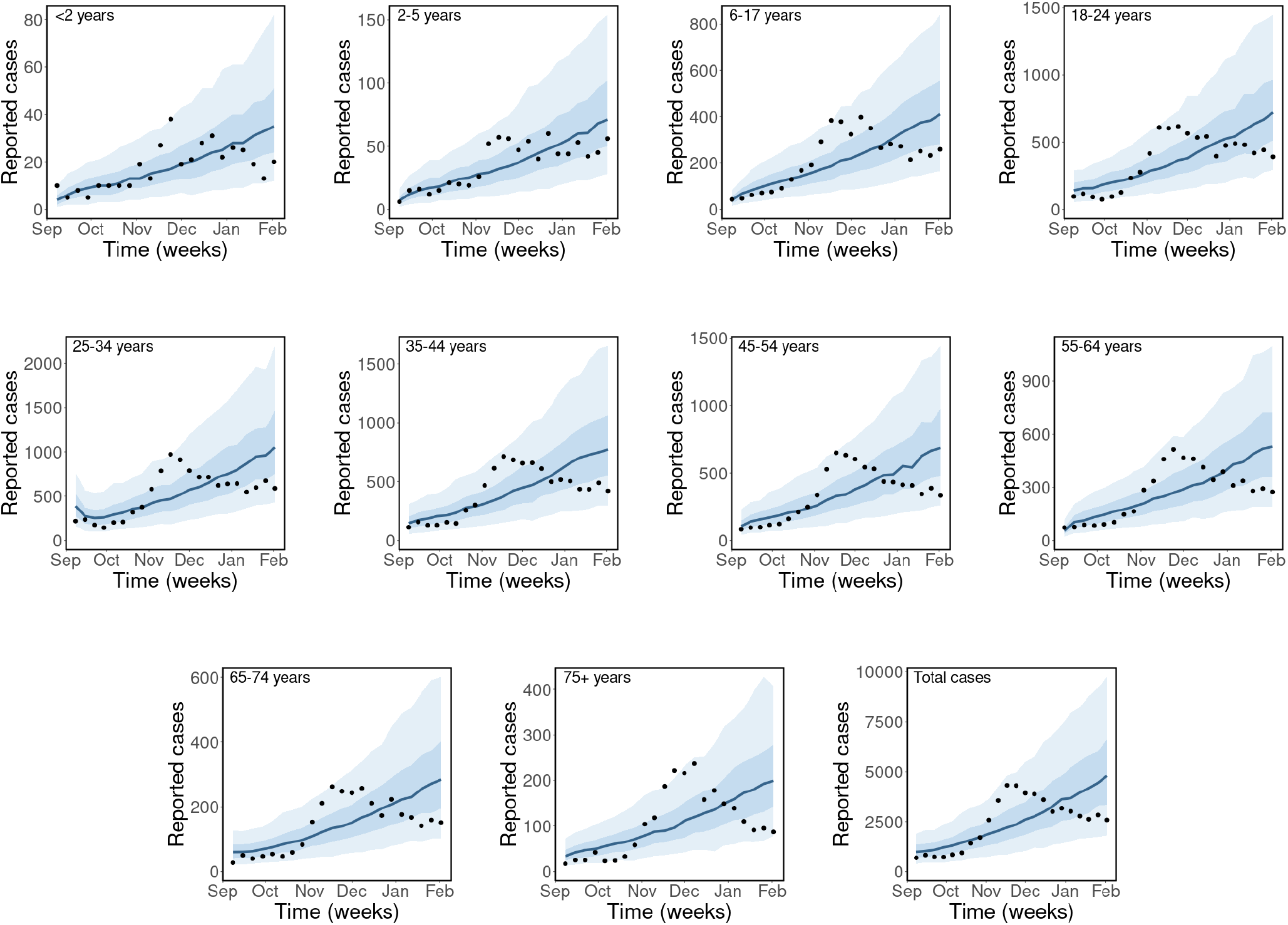
*Observed and estimated case counts by age group for the ASSFR model (age-specific scaling parameters and fixed contact rates) for the 2009 matrix. The black dots are the weekly reported cases of COVID-19 in BC, the blue solid lines are the median predicted cases, the narrower bands are the* 50% *CrI, while the wider bands are the 90*% *CrI*.

**Figure S8:**
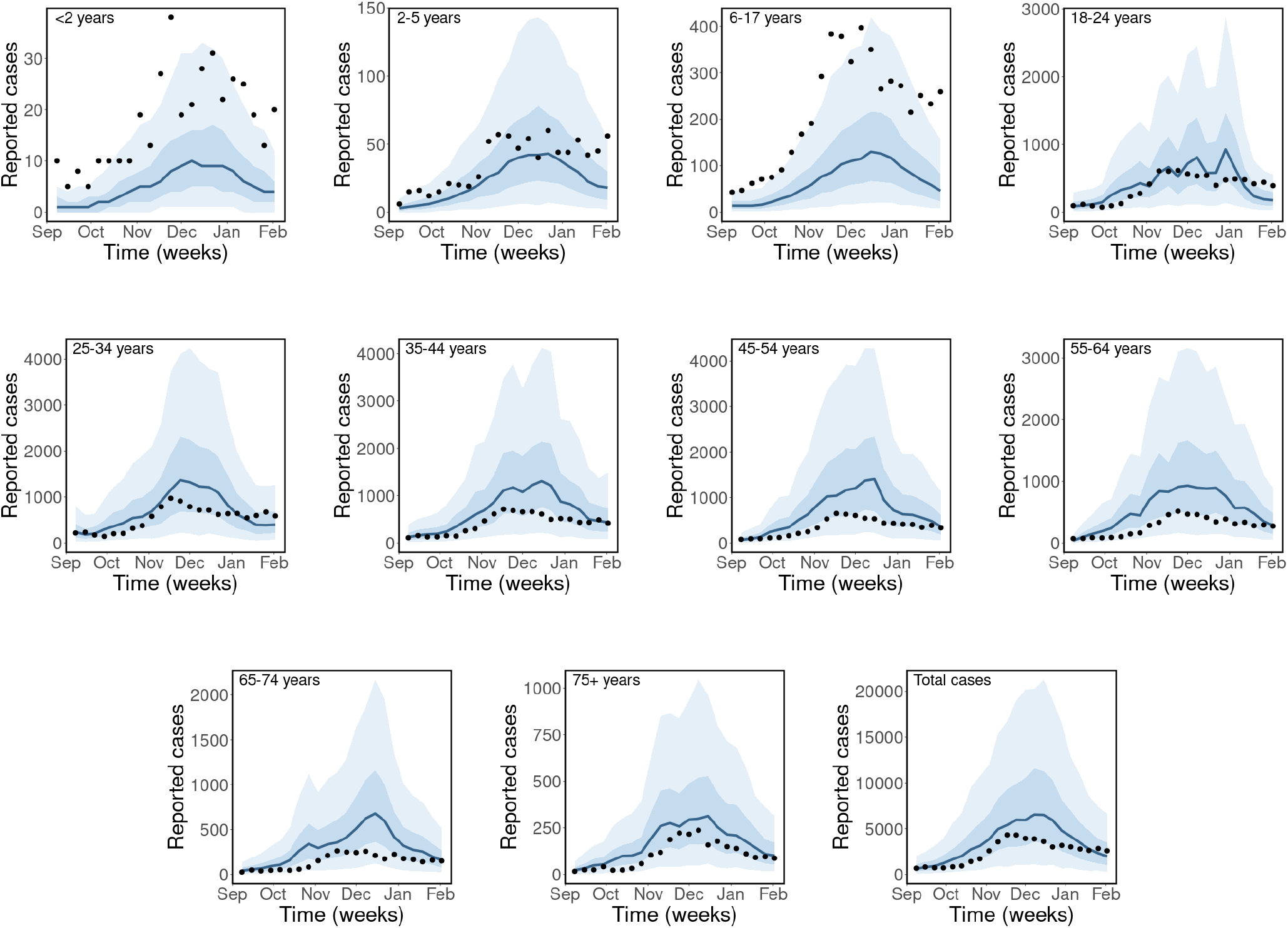
*Observed and estimated case counts by age group for the SSVR model (single scaling parameter and time-dependent weekly contact rates) for the 2009 matrix. The black dots are the weekly reported cases of COVID-19 in BC, the blue solid lines are the median predicted cases, the narrower bands are the* 50% *CrI, while the wider bands are the 90*% *CrI*.

**Figure S9:**
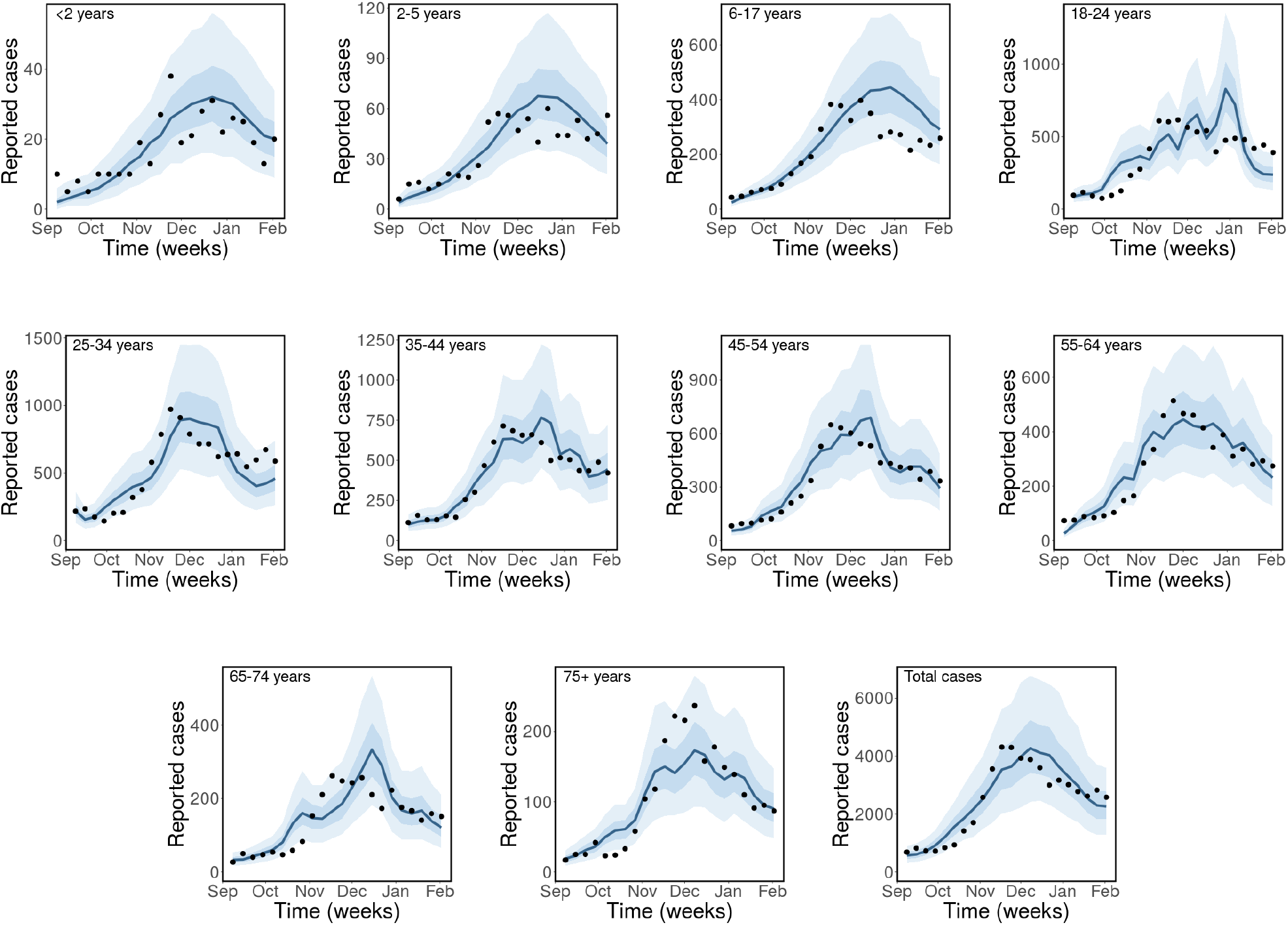
*Observed and estimated case counts by age group for the ASSVR scenario (age-specific scaling parameters and time-dependent average weekly contact rates) for the 2009 matrix. The black dots are the weekly reported cases of COVID-19 in BC, the blue solid lines are the median predicted cases, the narrower bands are the* 50% *CrI, while the wider bands are the 90*% *CrI*.

**Figure S10:**
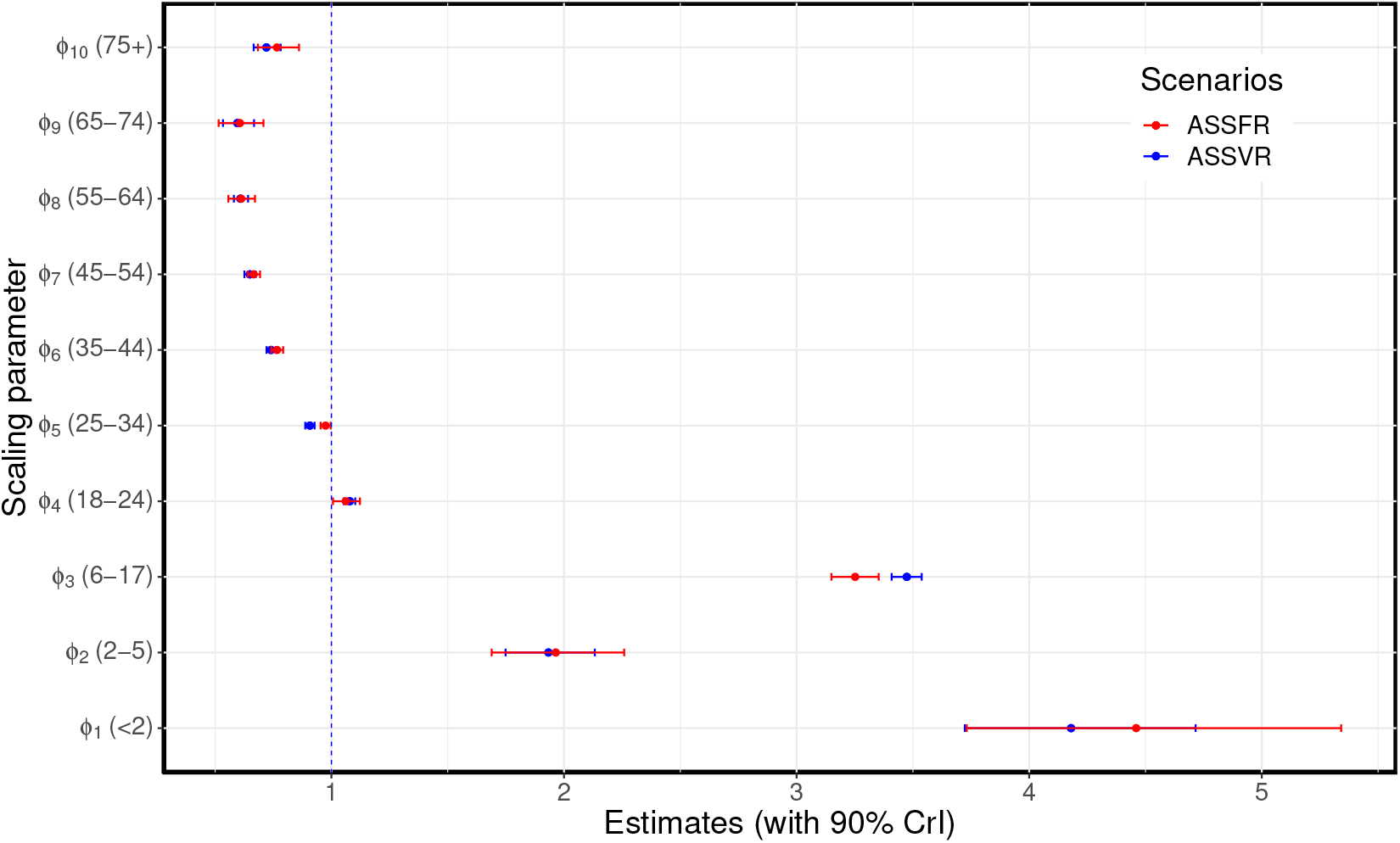
Estimated age-specific scaling parameters for the 2009 matrix. The ASSFR model (red) and ASSVR model (blue). The blue vertical dashed line at 1 indicates, where the scaling parameter has no effect on the fit. These estimates are presented in Tables S3.

**Table S3:**
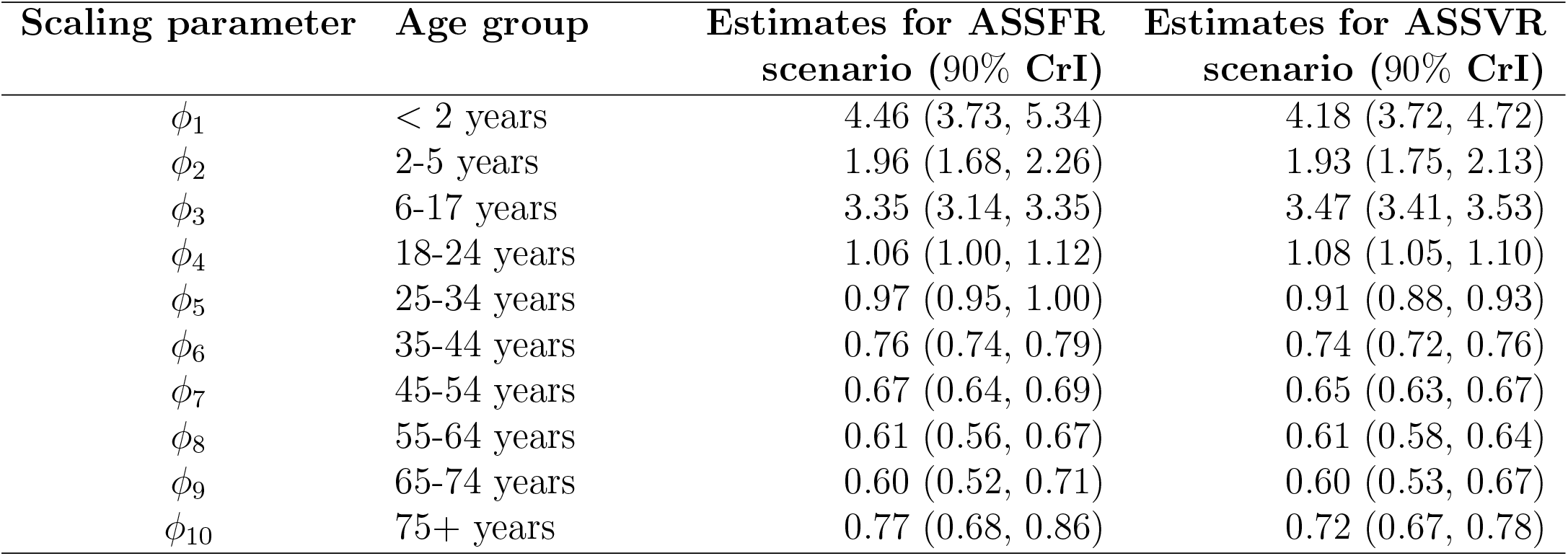
Estimated age-specific scaling parameters for the ASSFR and ASSVR models using the 2009 matrix.

